# Genome-wide association studies of 27 accelerometry-derived physical activity measurements identified novel loci and genetic mechanisms

**DOI:** 10.1101/2021.02.15.21251499

**Authors:** Guanghao Qi, Diptavo Dutta, Andrew Leroux, Debashree Ray, John Muschelli, Ciprian Crainiceanu, Nilanjan Chatterjee

## Abstract

Physical inactivity (PA) is an important risk factor for a wide range of diseases. Previous genome-wide association studies (GWAS), based on self-reported data or a small number of phenotypes derived from accelerometry, have identified a limited number of genetic loci associated with habitual PA and provided evidence for involvement of central nervous system in mediating genetic effects. In this study, we derived 27 PA phenotypes from wrist accelerometry data obtained from 88,411 UK Biobank study participants. Single-variant association analysis based on mixed-effects models and transcriptome-wide association studies (TWAS) together identified 5 novel loci that were not detected by previous studies of PA, sleep duration and self-reported chronotype. For both novel and previously known loci, we discovered associations with novel phenotypes including active-to-sedentary transition probability, light-intensity PA, activity during different times of the day and proxy phenotypes to sleep and circadian patterns. Follow-up studies including TWAS, colocalization, tissue-specific heritability enrichment, gene-set enrichment and genetic correlation analyses indicated the role of the blood and immune system in modulating the genetic effects and a secondary role of the digestive and endocrine systems. Our findings provided important insights into the genetic architecture of PA and its underlying mechanisms.

## Introduction

Regular physical activity (PA) is associated with lower risk of a wide range of diseases, including cancer, diabetes, cardiovascular disease(Kyu et al., 2016), Alzheimer’s disease(Rovio et al., 2005), as well as mortality(Smirnova et al., 2019; Leroux et al., 2019). However, studies have indicated that large majority of US adults and adolescents are insufficiently active(Piercy et al., 2018), and thus PA interventions have great potential to improve public health. PA was shown to have a substantial genetic component, and understanding its genetic mechanism can inform the design of individualized interventions(Lightfoot et al., 2018; Moore-Harrison & Lightfoot, 2010). For example, people who are genetically pre-disposed to low PA may benefit more from early and more frequent guidance.

A number of previous genome-wide association studies (GWAS) on physical activity have relied on self-reported phenotypes, which are subject to perception and recall error(De Moor et al., 2009; Hara et al., 2018; Kim et al., 2014; Klimentidis et al., 2018). Recently, wearable devices have been used extensively to collect physical activity data objectively and continuously for multiple days. To date, there have been two GWAS based on acceleromtery-derived activity phenotypes. Both studies used data from the UK Biobank study(Doherty et al., 2017; Bycroft et al., 2018) but only focused on a few summaries of these high-density PA measurements. One study considered two accelerometry-derived phenotypes (average acceleration and fraction accelerations > 425 milli-gravities) and identified 3 loci associated with PA(Klimentidis et al., 2018). A second study used a machine learning approach to extract PA phenotypes, including overall activity, sleep duration, sedentary time, walking and moderate intensity activity(Doherty et al., 2018). This study identified 14 loci associated with PA and found that the central nervous system (CNS) plays an essential role in modulating the genetic effects on PA. However, both studies used a small number of phenotypes, which may not capture the complexity of PA patterns.

Recent studies suggest that in addition to the total volume of activity, other PA summaries may be strongly associated with human health and mortality risk. For example, the transition between active and sedentary states was strongly associated with measures of health and mortality(Leroux et al., 2019; Schrack et al., 2019). PA relative amplitude, a proxy for sleep quality and circadian rhythm, was strongly associated with mental health(Rock, Goodwin, Harmer, & Wulff, 2014a). Moderate-to-vigorous PA (MVPA) and light intensity PA (LIPA) have also been reported to be associated with health(McGregor, Palarea-Albaladejo, Dall, Stamatakis, & Chastin, 2019; Young & Haskell, 2018). Thus, there is increasing evidence that objectively measured PA in the free-living environment is a highly complex phenotype that requires a large number of summaries that provide complementary information. Understanding the genetic mechanisms behind these summaries is critical for understanding the genetic regulation of activity behavior and informing targeted interventions.

In this paper, we conducted genome-wide association analysis using 27 accelerometry-derived PA measurements from UK Biobank data(Bycroft et al., 2018; Doherty et al., 2017). The phenotypes cover a wide range of features including volumes of activity, activity during different times of the day, active to sedentary transition probabilities, principal components and proxies for circadian rhythm (**Table 1**). We conducted GWAS using a mixed-model-based method, fastGWA(Jiang et al., 2019), to identify variants associated with the above phenotypes. We also conducted transcriptome-wide association studies (TWAS)(Gusev et al., 2016) across 48 tissues to identify genes and tissues harboring the associations. We further conducted colocalization (Giambartolomei et al., 2014), tissue-specific heritability enrichment(Finucane et al., 2018; Finucane et al., 2015), gene-set enrichment(Watanabe, Taskesen, van Bochoven, & Posthuma, 2017) and genetic correlation(Bulik-Sullivan et al., 2015a) analyses to further reveal the underlying biological mechanisms. We identified 5 novel loci associated with PA and showed that, in addition to the CNS, blood and immune related mechanisms could play an important role in modulating the genetic effects on activity, and digestive and endocrine tissues could play a secondary role.

**Table 1.**
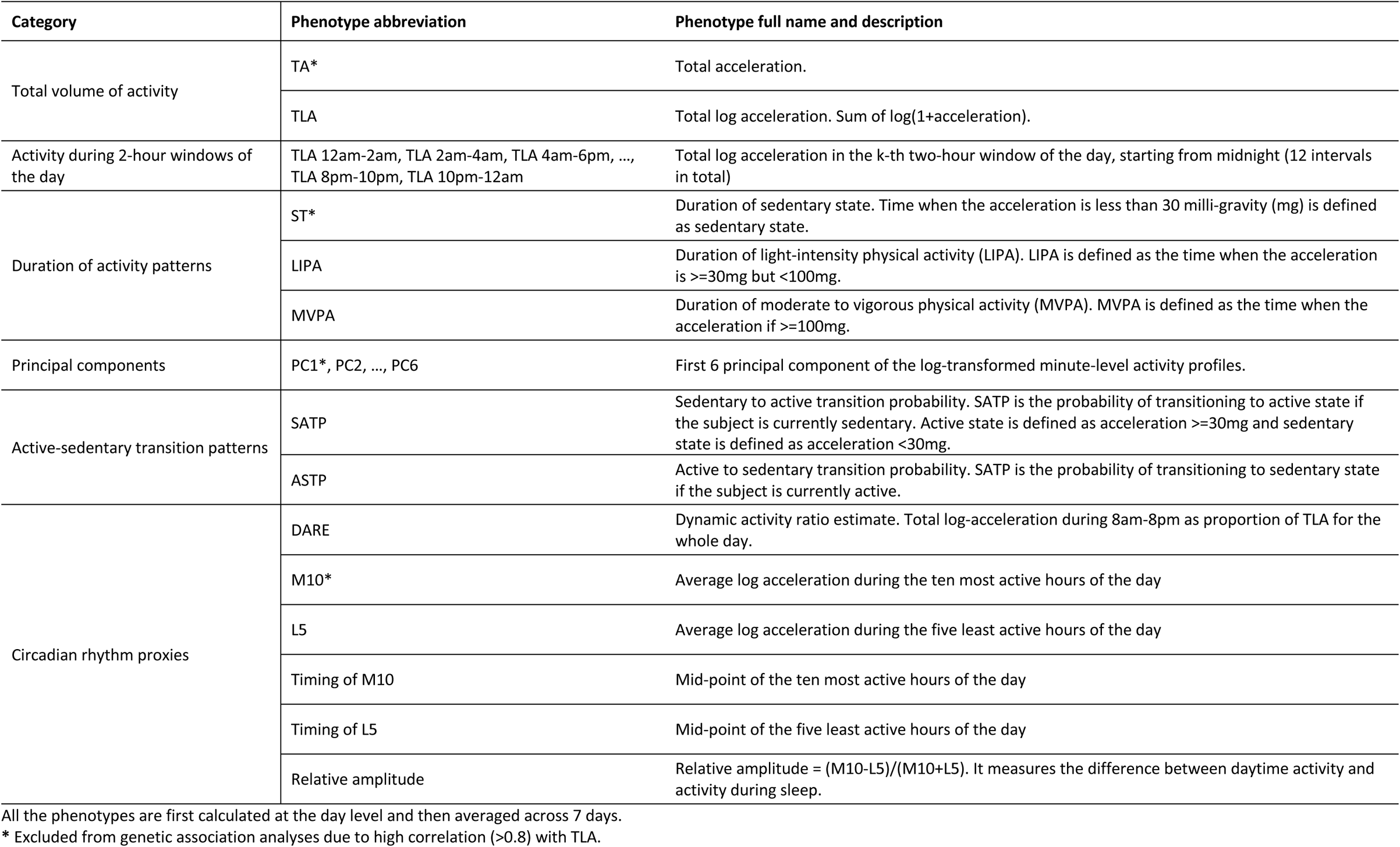
Description of physical activity phenotypes.

## Material and Methods

### Study Cohort and Physical Activity Phenotypes

The UK Biobank study consists of ~500,000 individuals in the United Kingdom with comprehensive genotype and phenotype data(Bycroft et al., 2018). We used a subset of the 103,712 individuals who were invited and agreed to participate in the accelerometry sub-study where participants wore a wrist-worn accelerometer for up to 7 days(Doherty et al., 2017).

Accelerometry data from participants are available at multiple resolutions. Here, the individual-specific set of accelerometry-based phenotypes was derived from the five-second level acceleration data provided by the UK Biobank team. We further compressed the data by averaging the 5-second level data within every minute. Individuals were screened for poor quality data using indictors provided by the UK Biobank. In addition, we required individuals to have at least 3 days (12am-12am) of sufficient wear time defined as estimated wear time greater than 95% of the day (>= 1368 minutes). Our inclusion criteria for this analysis closely mirrors that described in a related paper from our group(Leroux et al., 2020) with the exception that we did not exclude participants younger than 50 at the time of accelerometer wear or based on missing demographic and lifestyle data and instead excluded individuals based on ancestry and genotype data (see subsection *Genotype Data* below).

Physical activity phenotypes were all calculated at the day level and then averaged within study participants across days to obtain one measure for each phenotype and study participant. This led to in 31 PA phenotypes for 93,745 study participants that covered a wide spectrum of information. The phenotypes are described in **Table 1** and briefly summarizes as follows: 1) total volume of activity (total acceleration (TA), total log acceleration (TLA)); 2) activity during 12 disjoint two-hour windows of the day (TLA 12am-2am, TLA 2am-4am, …, TLA 10pm-12am); 3) duration of sedentary state (ST), LIPA and MVPA; 4) principal components of the log-transformed minute-level activity profiles (PC1-6); 5) active-to-sedentary transition probability (ASTP) and sedentary-to-active transition probability (SATP); 6) proxy phenotypes for circadian patterns, including dynamic activity ratio estimate (DARE), activity during the most active 10 hours (M10) and least active 5 hours (L5) of the day, timing of M10 and L5, and PA relative amplitude 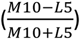. They included most of the phenotypes used in the previous PA association studies as well (See **Table 1** for details)(Doherty et al., 2018; Klimentidis et al., 2018). The exact procedure for deriving study participant-specific phenotypes is described in detail in the supplemental material of the related paper from our group(Leroux et al., 2020). The phenotypes were inverse-normal transformed

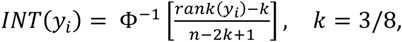

where the transformed variables have mean 0 and variance 1.

### Removing Highly Correlated Phenotypes

Some of the initial 31 PA phenotypes were highly correlated (**Figure S1**). To avoid counting similar phenotypes multiple times, if two phenotypes had correlation > 0.8 we removed one of them. First, we removed total acceleration (TA), duration of sedentary state (ST), PC1 and M10 due to their high correlation with total log acceleration (TLA). TLA was retained as the main metric for the total volume of activity. Since two previous GWASs used TA as the main metric for the volume of activity (Doherty et al., 2018; Klimentidis et al., 2018), we chose TLA instead of TA to avoid repetition with existing work. In addition, the original distribution of TA is highly skewed, which may not be completely addressed by the INT. In total, 4 phenotypes were removed and 27 PA phenotypes were retained for the association analysis.

### Genotype Data

The imputed genotype data for ~93 million variants, using UK10K, 1000 Genomes (Phase 3) and Haplotype Reference Consortium as reference panel, provided by UK Biobank were used and merged with the PA phenotype data. Following the Neale lab UK Biobank GWAS pipeline (https://github.com/Nealelab/UK_Biobank_GWAS/blob/master/imputed-v2-gwas/README.md), we excluded study participants according to the following criteria: 1) non-white ancestry; 2) putative sex chromosome aneuploidy; 3) an excessive number of relatives (more than 10 putative third-degree relatives in the kinship table); 4) sample was not in the input for phasing of chr1-chr22. After applying these exclusion criteria the sample was further reduced to 88,411 study particiants for downstream analysis.

We conducted variant quality control to ensure that genetic variants with poor genotyping quality do not affect the results. Specifically, variants that satisfy any of the following criteria were removed: 1) imputation INFO score < 0.8; 2) MAF < 0.01; 3) Hardy-Weinberg Equilibrium (HWE) p-value < 1 × 10^−6^; 4) missing in more than 10% study participants. After the filtering, 8,951,705 variants remained for downstream analysis, of which 8,067,228 (90.1%) were single nucleiotide polymorphisms (SNPs) and the rest (9.9%) were insertion-deletions (INDELs).

### Association Analysis

We used a fast mixed-effects model method, fastGWA(Jiang et al., 2019), for genome-wide association analysis. Like other mixed-effects model methods, fastGWA allows the inclusion of related and unrelated individuals but improves computational efficiency by incorporating a sparse genetic relationship matrix (GRM). The GRM measures the genetic similarity between individuals and each element is the correlation of genotypes between a pair of individuals. We constructed the GRM using linkage disequilibrium (LD)-pruned variants that had MAF > 5% and were present in HapMap3 (LD-pruning was done in PLINK using the following set up as recommended in Jiang et al (Jiang et al., 2019): window size = 1000Kb, step-size = 100 and r^2^ = 0.9). We further computed a sparse-GRM at sparsity level 0.05 to capture the genetic relatedness between the closely related individuals only and reduced others to zero. We used the Haseman-Elston regression to estimate the variance of the random effects as an intermediate step of fastGWA. This approach is orders of magnitude faster than the previous state-of-the-art, BOLT-LMM(Loh et al., 2015; Loh, Kichaev, Gazal, Schoech, & Price, 2018).

Models were adjusted for age, sex and the first 20 genetic principal components as covariates. Because the PA phenotypes are correlated, principal component analysis (PCA) was conducted on the phenotypes to estimate the number of independent phenotypes before setting the GWAS significance threshold. At least 19 phenotype PCs were needed to explain 99% percent of the PA phenotypic variance (**Figure S2**). Variants with p-value below the threshold 5 × 10^−8^/19 = 2.63 × 10^−9^ were declared to be statistically significant, which accounted for the number of independent phenotypes. LD clumping was conducted based on the minimum p-value across phenotypes. The requirements for the lead SNPs of different loci were to have *r*^2^< 0.1 and be at least > 500kb apart.

A locus was defined as novel if its lead variant is >500kb from the lead variant of any known loci discovered by the following GWASs on PA, sleep, and circadian rhythm: (1) Doherty et al study on a smaller set of accelerometry-derived PA phenotypes(Doherty et al., 2018); (2) Klimentidis et al study on self-reported and accelerometry-derived PA(Klimentidis et al., 2018); (3) Dashti et al study on self-reported sleep duration(Dashti et al., 2019); (4) Jones et al study on circadian rhythm(Jones et al., 2019). Considering the diversity of PA phenotypes, we further ensure these loci do not have associations with other related traits not listed above, by searching for the lead variants in Open Targets Genetics (OTG, https://genetics.opentargets.org/). A locus remains novel if the lead variant: 1) is not associated at *p* < 5 × 10^−8^ with traits whose names include the following keywords: accelerometry, physical, exercise, sleep, nap, circadian and chronotype; 2) and is not in LD with previously reported GWAS lead variants for these traits (*r*^2^ > 0.5).

Transcriptome-wide association studies (TWAS) were conducted using the FUSION R program(Gusev et al., 2016) with reference models generated from 48 tissues of GTEx v7(GTEx et al., 2017). TWAS analysis was limited to the 18 traits with at least one genome-wide significant variant (*p* < 2.63 × 10^−9^). Multiple testing due to the large number of tissue-trait combinations (48*18=864) was addressed by a two-stage adjustment approach: 1) for each variant, the Benjamini-Hochberg (BH) adjustment was applied across all tissue-trait pairs; 2) each variant with BH-adjusted p-value 2.5 × 10^−6^ was then identified (accounting for 20,000 protein-coding genes). Since there can be multiple genes in close proximity to each other, to identify independent loci detected by TWAS analysis, genes were clustered based on significant associations. A clumping approach was used, which selected the gene with the smallest minimum p-value across tissue-trait pairs and removed the other genes with a transcription start site (TSS) within 1Mb of the lead gene TSS. The process continued by identifying the gene with the next smallest minimum p-value and iterating. The only exception was when the lead gene of the cluster was not a protein-coding gene (e.g., pseudogene, lncRNA) and a protein-coding gene was in the cluster. In this case the protein-coding gene with the smallest minimum p-value was identified as the lead gene. This led to independent gene clusters at genomic loci which were least 1Mb apart, i.e., none of the lead gene TSS is within the cis region of another lead gene.

### Enrichment Analysis

Stratified LD score regression(Finucane et al., 2015; Finucane et al., 2018) was used to identify the tissues and genomic annotations enriched by the heritability for PA. For tissue specific analysis, chromatin-based annotations were used as derived from the ENCODE and Roadmap data(ENCODE, 2012; Roadmap et al., 2015) by Finucane et al(Finucane et al., 2018). The annotations were based on narrow peaks of DNase I hypersensitivity site (DHS) and five activating histone marks (H3K27ac, H3K4me3, H3K4me1, H3K9ac and H3K36me3) observed for 111 tissues or cell types, resulting in a total of 489 annotations. Stratified LD score regression computes the heritability attributed to each annotation and computes a coefficient and a p-value that characterize enrichment.

In a separate analysis, the enrichment of TWAS signals was evaluated among the genes that have been reported to be associated to different traits, using FUMA(Watanabe et al., 2017; Watanabe, Umićević Mirkov, de Leeuw, van den Heuvel, & Posthuma, 2019). For a given PA trait, we defined a gene-set as the genes that were significant at an exome-wide level (*p* < 2.5 × 10^−6^) and investigated whether these genes overlapped with the genes that have been mapped to genome-wide significant variants for different traits as reported in GWAS catalog(Buniello et al., 2019). The collection of such genes have been detailed in Molecular Signatures Database (MSigDB)(Liberzon et al., 2011). We used FUMA to compute the proportion of genes related to other diseases and traits that were also identified by our TWAS analysis and computed enrichment p-values using the Fisher’s exact test.

### Colocalization Analysis

For each susceptibility locus of PA (**Table 2**), colocalization analysis was conducted between its most significantly associated phenotype and eQTL effects on gene expression in 48 tissues in GTEx v7(GTEx et al., 2017). SNPs within +-200kb radius of the lead SNP were used and genes that had at least one significant eQTL (q-value < 0.05) in the region were considered. Analysis was conducted using the R package COLOC (Giambartolomei et al., 2014) and GWAS and eQTL effects were identified as being colocalized if PP4 > 0.8.

**Table 2.** Significant loci associated with physical activity in single-variant analysis.

| Lead variant<sup>a</sup> | Chromosome region | Base pair | Minor allele | Major allele | Minor allele frequency | Nearest coding gene and distance<sup>d</sup> | Significantly associated traits<sup>c</sup> | Previous studies that discovered the locus |
| --- | --- | --- | --- | --- | --- | --- | --- | --- |
| Novel loci<sup>b</sup> |  |  |  |  |  |  |  |  |
| rs3836464 | 3p25.3 | 10454772 | CA | C | 0.274 | SEC13 (92kb) | ASTP (p=2.2e-09, b=-0.032) | NA |
| rs9818758 | 3p21.31 | 49382925 | A | G | 0.171 | USP4 (4.8kb) | Relative amplitude (p=2.1e-09, b=-0.036) |  |
| 3:131647162_TA_T | 3q22 | 131647162 | T | TA | 0.466 | CPNE4 (357kb) | TLA 2am-4am (p=2.2e-09, b=0.029) |  |
| Known loci |  |  |  |  |  |  |  |  |
| rs1144566 | 1q25.3 | 182569626 | T | C | 0.030 | RGS16 (3.9kb) | Timing of L5 (p=5e-10, b=-0.086)* | 4 |
| rs301799 | 1p36.23 | 8489302 | C | T | 0.422 | SLC45A1 (111kb) | TLA 6pm-8pm (p=1.7e-09, b=0.027)* | OTG<sup>e</sup> |
| rs113851554 | 2p14 | 66750564 | T | G | 0.050 | MEIS1 (90kb) | TLA 12am-2am (p=6.7e-37, b=0.138)*, TLA 2am-4am (p=7.9e-39, b=0.142)*, L5 (p=1.3e-33, b=0.13)*, Relative amplitude (p=6.9e-15, b=-0.082)*, Timing of L5 (p=5.4e-22, b=0.105)* | 1,4 |
| rs2909950 | 5q33.1 | 151886147 | A | G | 0.418 | NMUR2 (73kb) | TLA 6pm-8pm (p=9.4e-10, b=-0.028)* | 4 |
| rs12717867 | 5q33.1 | 152412845 | G | A | 0.453 | GRIA1 (456kb) | LIPA (p=6.2e-10, b=-0.029)* | 4 |
| rs9369062 | 6p21.2 | 38437303 | C | A | 0.292 | BTBD9 (171kb) | TLA 12am-2am (p=1.5e-12, b=-0.037)*, TLA 2am-4am (p=2.3e-10, b=-0.033)* | 4 |
| rs2006810 | 7q11.22 | 69902152 | C | T | 0.395 | GALNT17 (695kb) | TLA 8pm-10pm (p=8.1e-11, b=-0.031)* | 1,4 |
| rs1268539 | 9q33.3 | 128195657 | A | C | 0.419 | GAPVD1 (172kb) | TLA (p=1.1e-10, b=0.03), LIPA (p=3.2e-10, b=0.03)* | 1 |
| rs564819152 | 10p12.31 | 21820650 | G | A | 0.320 | SKIDA1 (5kb) | TLA 8am-10am (p=1.4e-09, b=-0.031)* | 1,3 |
| rs2138543 | 12q12 | 39298423 | A | T | 0.477 | CPNE8 (2.8kb) | TLA 6am-8am (p=3.9e-11, b=0.03)*, PC2 (p=1.5e-09, b=-0.029)* | 4 |
| rs12927162 | 16q12.2 | 52684916 | G | A | 0.277 | TOX3 (103kb) | TLA 10pm-12am (p=1e-09, b=0.032)* | 4 |
| rs2532402 | 17q21.31 | 44304130 | G | C | 0.221 | KANSL1 (1.4kb) | TLA (p=1.5e-12, b=0.04), MVPA (p=1.9e-10, b=0.035) | 1,2,3,4 |
| rs3837946 | 19p13.2 | 9955920 | TTTTG | T | 0.475 | PIN1 (10kb) | TLA (p=1.3e-11, b=-0.032), LIPA (p=1.6e-09, b=-0.029)* | 1,3 |

<sup>a</sup>Significant variants (single nucleotide polymorphism (SNP) or insertion/deletion) are defined as those with fastGWA p-value < $2.63 \times 10^{-9}$ (Bonferroni corrected for 19 independent traits). LD clumping was performed at $r^2 < 0.1$ and lead variants of different loci were required to be >500kb apart.
<sup>b</sup>A locus is defined as novel if its lead variant is >500kb from the lead variant of any loci identified in one of the previous GWAS: 1) Doherty et al study on a smaller set of accelerometry-based physical activity; 2) Klimentidis et al study on self-reported and accelerometry based physical activity; 3) Dashti et al study on self-reported sleep duration; 4) Jones et al study on circadian rhythm. Loci that were discovered by the four above studies are marked with 1, 2, 3 and 4 in the last column, respectively. Novel phenotypes associated with known loci are marked with \*.
<sup>c</sup>Traits are followed by p-values (p) and effect sizes (b). The effects size b is for the effect of minor allele vs major allele, e.g. a positive b indicates the minor allele is the trait-increasing allele. TLA: total log-acceleration. ASTP: active-to-sedentary transition probability. MVPA: moderate-to-vigorous physical activity. PC2: second principal component. LIPA: light-intensity physical activity.
<sup>d</sup>Nearest coding gene is defined as the protein-coding gene whose transcription start site (TSS) is closest to the variant.
<sup>e</sup>This locus was not reported by the four studies we catalogued but was associated with daytime nap, as reported by Open Targets Genetics (OTG). See Table S4 for details.

### Heritability and Genetic Correlation Analysis

Heritability of activity phenotypes was estimated using Haseman-Elston regression as an intermediate output of fastGWA(Jiang et al., 2019). Our fastGWA analysis computed sparse GRM at sparsity level 0.05 as recommended by the fastGWA paper (see “Association analysis”). However, this cutoff may miss the subtle relatedness in the sample and affect heritability estimate. As a sensitivity analysis, we re-estimated the heritability using a lower sparsity threshold at 0.02 to capture more subtle relatedness. The genetic correlation between 18 PA traits and 238 complex traits and diseases was estimated using LD score regression(Bulik-Sullivan et al., 2015a) implemented in LD Hub(Zheng et al., 2017). In particular, we focused on four broad groups of traits and diseases (A) cholesterol levels (B) anthropometric traits (C) autoimmune disease and (D) miscellaneous traits including psychiatric, neurological, cognitive and personality traits. For each trait and within each category, we applied a false discovery rate correction to the p-values corresponding to the genetic correlation estimated using LD score regression, to account for multiple testing. Any genetic correlation with FDR-adjusted p-value less that 10% were declared as significant.

## Results

### Genetic Loci Associated with Physical Activity

Single-variant genome-wide association analysis identified a total of 16 independent loci, including three novel ones compared to previous studies (**Table 2 and Figure 1**). All three novel loci were discovered on chromosome 3: the locus indexed by rs3836464 was associated with ASTP; the locus indexed by rs9818758 was associated with relative amplitude, which is a proxy sleep behavior and circadian rhythm(Rock, Goodwin, Harmer, & Wulff, 2014b); the locus indexed by indel 3:131647162_TA_T (no rsid available) was associated with TLA 2am-4am which is a proxy phenotype for activity during sleep. LIPA appeared to be associated with other SNPs near 3:131647162_TA_T but not the lead variant itself, indicating multiple independent signals at the same locus (**Figure 1**). Nearest coding genes for the novel loci include *SEC13*, *USP4*, and *CPNE4*.

**Figure 1.**
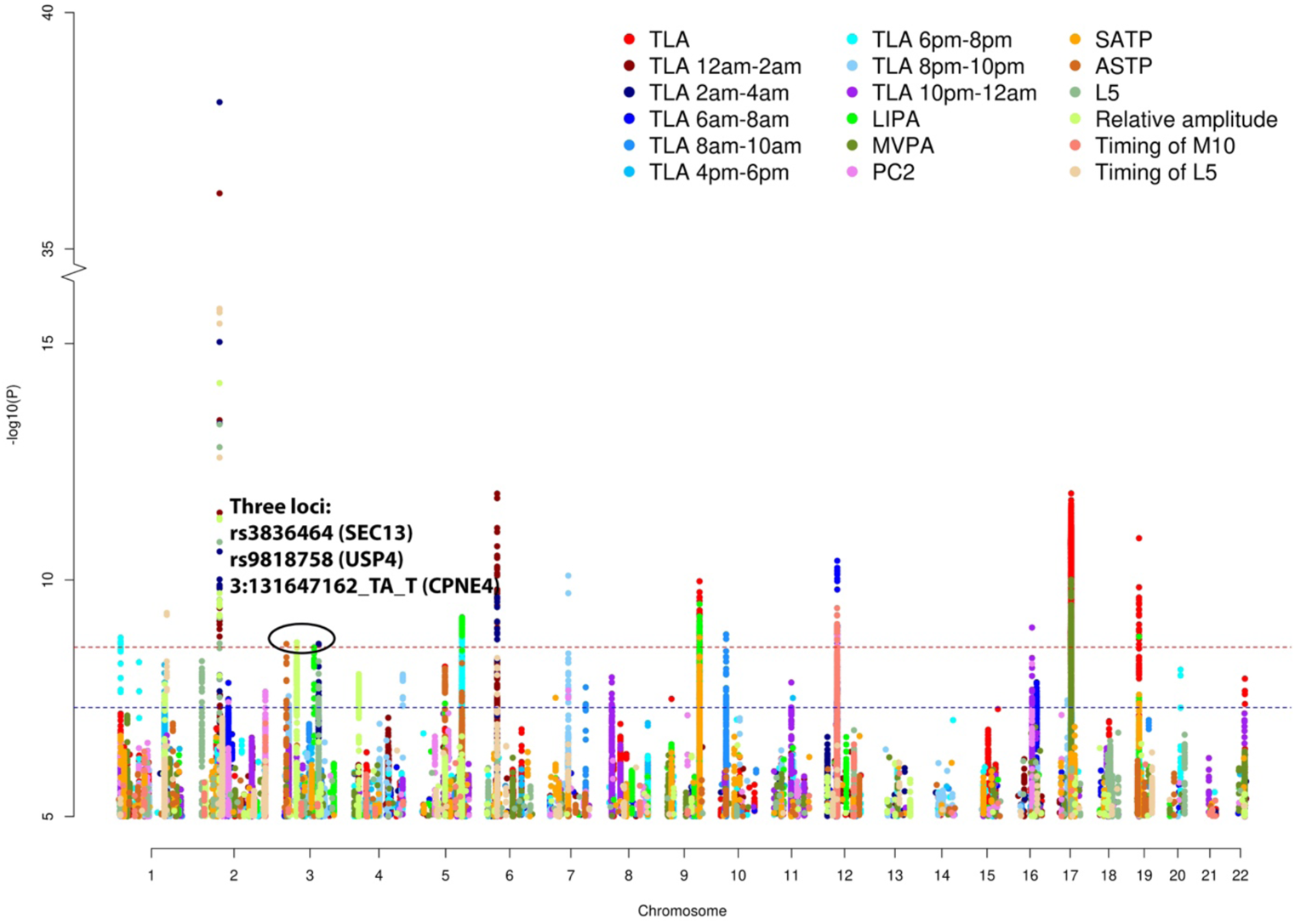
Manhattan plot for 18 traits that are significantly associated with at least one variant at *p* <2.63 × 10^−9^ **in single-variant analysis.** The red dashed line is *p* = 2.63 × 10^−9^ accounting for the number of independent traits. The blue dashed line is the standard genome-wide significance threshold *p* = 5 × 10^−8^. Three novel loci that have not been discovered in previous GWAS of physical activity, sleep duration and circadian rhythm are circled out and annotated by the lead variant and nearest coding gene (see Table 2).

Our analysis also identified novel phenotypes for several known loci (**Table 2**). The strongest signal was seen for the locus indexed by rs113851554 which is associated with multiple sleep and circadian rhythm proxy phenotypes including TLA 12am-2am (*p* = 6.7 × 10^−37^), TLA 2am-4am (*p* = 7.9 × 10^−39^), average log acceleration during the least active 5 hours of the day (L5, *p* = 1.3 × 10^−33^), timing of L5 (*p* = 5.4 × 10^−22^) and PA relative amplitude (*p* = 6.9 × 10^−15^). This locus was previously identified to be associated with accelerometry-derived sleep duration in UK Biobank(Doherty et al., 2018). Among other known loci, 5 were only discovered in the GWAS of self-reported circadian rhythm(Jones et al., 2019) but not in the other studies considered (**Table 2**, last column). In our analysis, the loci indexed by rs1144566, rs9369062 and rs12927162 were associated with sleep proxy phenotypes including timing of L5, TLA 12am-2am, TLA 2am-4am and TLA 10pm-12am. Three other loci, indexed by rs301799, rs2909950 and rs12717867, were associated with TLA 6pm-8pm and LIPA, respectively.

In addition to the phenotypic associations above, other variants in some of the loci captured associations that are not reflected by the lead variants. In particular, variants in high LD with rs2138543 are associated with a wide range of phenotypes including LIPA, MVPA, activity during two-hour windows, and a number of proxy phenotypes for circadian patterns (**Table S1**).

### Transcriptome-Wide Association Study and Colocalization Analysis

We performed transcriptome-wide association studies (TWAS) (Gamazon et al., 2015; Gusev et al., 2016) for each PA trait based on gene expression data across 48 tissues available through GTEx (version 7)(GTEx et al., 2017). Our analysis identified 15 loci (**Table 3, Figure 2, Table S2**) with significant association in at least one trait-tissue pair analysis after correcting for multiple testing (Benjamini-Hochberg corrected p-value < 2.5 × 10^−6^, see **Methods**). We identified two novel loci. One of them was indexed by *RN7SKP16*, whose higher expression in brain putamen basal ganglia is genetically associated with lower level of MVPA. Another was indexed by pseudogene *PDXDC2P (16q22.1)*, whose higher expression in esophagus mucosa and EBV transformed lymphocytes appeared to be genetically associated with lower level of TLA 6am-8am (**Figure 2**). See **Table S2** for details of these associations. These loci was not previously reported by any prior GWAS and were not close to any of the 3 novel regions detected by our single variants analysis (**Table 3**).

**Figure 2:**
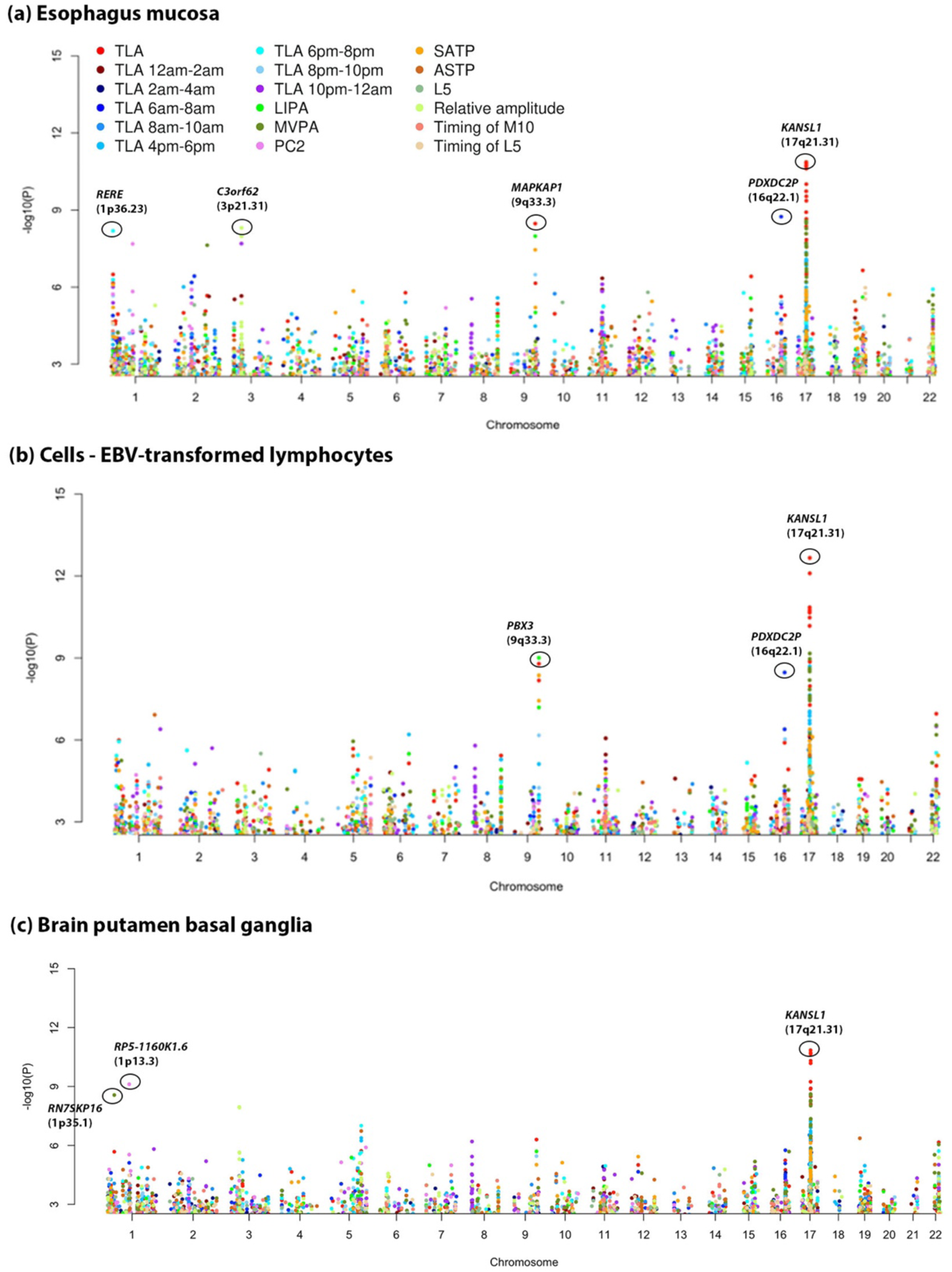
**TWAS Manhattan plots for three tissues that harbor the TWAS novel loci**. Significant genes with FDR corrected p-values< 2.5 × 10^−6^ are circled (see Methods for details). Only the PA traits that are significantly associated with at least one variant at *p* < 2.63 × 10^−9^ in single-variant analysis are shown.

**Table 3.** Significant loci associated with physical activity identified in transcriptome-wide association studies (TWAS).

| Most significant gene<sup>a</sup> | CHR | Gene start | Gene end | Locus | Minimum p-value across traits and tissues | Secondary genes in the locus with comparable associations<sup>c</sup> | Trait and tissue<sup>d</sup> | Previous studies that discovered the locus |
| --- | --- | --- | --- | --- | --- | --- | --- | --- |
| Novel loci<sup>b</sup> |  |  |  |  |  |  |  |  |
| *RN7SKP16* | 1 | 33802167 | 33802465 | 1p35.1 | 2.72E-09 | - | MVPA (CNS) |  |
| *PDXDC2P* | 16 | 70069541 | 70098679 | 16q22.1 | 1.81E-09 | - | TLA 6am-8am (Blood/Immune, Digestive) | NA |
| Known loci |  |  |  |  |  |  |  |  |
| *RERE*<sup>†</sup> | 1 | 8412457 | 8877702 | 1p36.23 | 5.28E-09 | *ENO1-IT1, RP5-1115A15.1*<sup>†</sup> | TLA 6pm-8pm (Digestive, Other, Blood/Immune) | GWAS |
| *RP5-1160K1.6* | 1 | 110171118 | 110171939 | 1p13.3 | 7.68E-10 | - | PC2 (CNS) | 4 |
| *C3orf62*<sup>†</sup> | 3 | 49306219 | 49315263 | 3p21.31 | 3.28E-10 | *RP11-694I15.7, ARIH2, DAG1, SLC25A20* | Relative amplitude (Digestive, Endocrine) | GWAS |
| *CTC-467M3.3* | 5 | 87988462 | 87989789 | 5q14.3 | 1.39E-08 | - | ASTP (CNS) | 1,4 |
| *NMUR2* | 5 | 151771093 | 151812929 | 5q33.1 | 8.88E-10 | - | TLA 6pm-8pm (CNS, Other) | 4, GWAS |
| *PBX3*<sup>†</sup> | 9 | 128509624 | 128729656 | 9q33.3 | 3.04E-10 | *MAPKAP1* | LIPA (Blood/Immune, Digestive, Endocrine, Other); SATP (Blood/Immune, Other, Digestive, Endocrine); TLA (Blood/Immune, Other, Digestive, Endocrine) | 1, GWAS |
| *DNAJC1* | 10 | 22045466 | 22292698 | 10p12.31 | 3.20E-09 | *CASC10*<sup>†</sup> | TLA 8am-10am (CNS) | 1,3, GWAS |
| *RP11-396F22.1*<sup>†</sup> | 12 | 39300253 | 39303394 | 12q12 | 9.89E-10 | - | Timing of M10 (CNS), PC2 (CNS, Other), TLA 6am-8am (CNS, Other) | 4, GWAS |
| *FMNL1* | 17 | 43299590 | 43324633 | 17q21.31 | 2.82E-09 | *CTD-2020K17.1* | TLA (CNS) | 1,4 |
| *KANSL1* | 17 | 44107282 | 44302733 | 17q21.31 | 2.17E-13 | *KANSL1-AS1, RP11-798G7.8* | MVPA (CNS, Blood/Immune, Other, Digestive, Musculoskeletal/connective); TLA 4pm-6pm (Blood/Immune, Other, Digestive); TLA (Adipose, CNS, Blood/Immune, Other, Digestive, Cardiovascular, Musculoskeletal/connective, Endocrine) | 1,2,3,4, GWAS |
| *ZNF846* | 19 | 9862669 | 9903856 | 19p13.2 | 2.22E-10 | *OLFM2, PIN1, CTD-2623N2.3* | TLA (Other) | 1,3, GWAS |
| *JUND* | 19 | 18390563 | 18392432 | 19p13.11 | 3.18E-09 | - | SATP (Other) | 4 |
| *LINC00634* | 22 | 42348169 | 42354937 | 22q13.2 | 4.85E-11 | - | TLA (CNS) | 4 |

<sup>a</sup>For each gene, we apply Benjamini-Hochberg (BH) correction across all trait-tissue pairs. Significant genes are defined as those with BH corrected p-value < $2.5 \times 10^{-6}$ . Significant genes are clustered using an approach similar to LD clumping so that different loci, marked by the transcription start site (TSS) of the lead gene, are >1Mb apart. Gene symbols are italicized. <sup>†</sup> Genes of which the eQTL signal colocalizes with GWAS signal of the most significantly associated phenotype.
<sup>b</sup>A locus is defined as novel if its flanking region ( $\pm 500$ kb from TSS) does not harbor a variant reported in Table 2 or any of the four previous studies considered in Table 2. Loci that were discovered by the above studies are marked with 1, 2, 3 and 4 in the last column, respectively. Loci that were discovered by our single-variant analysis (Table 2) are marked with "GWAS" in the first column.
<sup>c</sup>Other genes in the cluster with minimum p-value less than 10 times the minimum p-value of the lead gene are shown in column "Secondary genes in the locus with comparable associations". See Table S2 for all the significant tissue-trait pairs.
<sup>d</sup>CNS: central nervous system. TLA: total log-acceleration. ASTP: active-to-sedentary transition probability. SATP: sedentary-to-active transition probability. MVPA: moderate-to-vigorous physical activity. PC2: second principal component.

The TWAS analysis also identified novel PA phenotypes, potential target genes and underlying tissues for many of the known loci or novel loci detected through single variant analysis (**Table 3**). Consistent with a previous study (Doherty et al., 2018), the TWAS analysis showed that genetic association for PA traits often points towards involvement of CNS (**Table 3**). Further, our analysis indicates consistent involvement of blood and immune, digestive and endocrine systems in modulating the genetic effects on PA. See **Table S2** for a complete list of associated tissues. Among the 15 loci significant in TWAS analysis, the lead genes of 4 loci were significantly associated with PA phenotypes via the blood and immune tissues. For example, the genetically predicted expression of *PBX3* and *KANSL1* in the blood and immune tissues (whole blood, EBV-transformed lymphocytes) were each associated with 3 PA phenotypes. The genes associated with PA via blood and immune tissues were also associated via digestive (esophagus mucosa, small intestine - terminal ileum, colon – sigmoid, etc) and/or endocrine (thyroid, pituitary) tissues, but only one of them overlapped with the 9 genes that were associated with PA phenotypes through the CNS (**Table 3**). Another locus, represented by *C3orf62*, were associated with PA relative amplitude only via the digestive and endocrine tissues (esophagus mucosa, colon – sigmoid, thyroid) but not the blood/immune tissues or CNS. These findings suggested two potential pathways for the genetic regulation of PA: a primary pathway involving the CNS (brain in particular) and a secondary pathway involving the blood/immune system and, potentially, the digestive and endocrine systems. The actual biological processes involved in the pathways are beyond the scope of this paper and may be worth future investigation.

Several genes that were found to be significantly associated to specific PA traits in our TWAS analysis, were also found to be highly overlapping with genes that were previously reported to be associated with various traits and diseases including but not limited to neuropsychiatric diseases, behavioral traits, anthropometric traits and autoimmune diseases (**Figure S3**). For example, we found that the genes associated with TLA across different tissues, are enriched for genes that have been associated with neuroticism, bipolar disorder, Parkinson’s disease, cognitive function and several others indicating the putative involvement of the CNS in the genetic mechanism of TLA. Additionally, the genes associated with relative amplitude overlapped highly with those associated with several autoimmune diseases like inflammatory bowel disease, ulcerative colitis in addition to different behavioral and cognitive traits (**Figure S3**). These results further supported the possible involvement of both CNS as well as the blood and immune system in the genetic mechanism of PA traits.

We performed a colocalization analysis to gain further insights on the tissue specific activity of the significant genetic loci. Among the 16 loci significantly associated with PA, 9 loci colocalized with the eQTL signals for at least one gene and one tissue with a colocalization probability (PP4) > 0.8 (**Table S3**). Colocalization occurred in a similar set of tissues as those that harbored the TWAS associations (**Table 3 and Table S3**), namely the CNS, blood and immune (whole blood, spleen, EBV transformed lymphocytes), digestive (esophagus, colon) and endocrine (thyroid, testis, adrenal gland) tissues, and also in a number of cardiovascular tissues that were not highlighted by TWAS. Among the 15 lead genes for TWAS significant loci, the eQTL signal of 4 genes (*RERE, C3orf62, PBX3* and *RP11-396F22.1*) colocalized with PA GWAS signal in at least one tissue. Colocalization also occurred in two other secondary genes (*RP5-1115A15.1* and *CASC10*).

### Analysis of Heritability and Co-Heritability

Our fastGWA analysis estimated genome-wide heritability of PA phenotypes as an intermediate output. The estimates appeared to be dependent on the sparsity level of the genetic relationship matrix (**Figure S4**). We chose the results under the lower cutoff (0.02) since it captured more subtle relatedness and should give more accurate heritability estimates. The estimates of heritability varied across different PA phenotypes. A number of traits were estimated to have higher heritability than others, including TLA (0.15), TLA 6pm-8pm (0.15), MVPA (0.14) (**Figure S4**). Afternoon and pre-sleep evening activity (TLA 4pm to 12am) appeared to be more heritable than morning activity (TLA 2am to 12pm). As could be expected, phenotypes with higher heritability tend to have a higher average @^0^ statistic for genetic associations, and a QQ plot which deviate further from the null line (**Figure S5**). The magnitude of heritability estimates was generally consistent with previous studies, which reported 10-20% heritability for PA traits (Doherty et al., 2018; Klimentidis et al., 2018). We also notice that heritability estimated using restricted maximum likelihood (REML) tended to be slightly higher Haseman-Elston regression estimates (**Figure S4**).

We further used stratified LD-score regression for partitioning heritability by functional annotations of genome(Finucane et al., 2015; Finucane et al., 2018). Consistent with TWAS findings, this analysis also indicated possible role for blood and immune system in addition to CNS for genetic regulation of PA (**Figure 3**). In particular, heritabilities for both TLA and LIPA were enriched for DNase I hypersensitivity sites (DHS) in primary B cells from peripheral blood and that for TLA 12pm-2pm were enriched for H3K27ac in spleen. We also found potential enrichment in other traits, though they were not significant after FDR adjustment. For example, for TLA 8am-10am, MVPA, and ASTP the heritability enrichment in active chromatin regions of blood/immune tissues were all close to being statistically significant (**Figure S6**).

**Figure 3.**
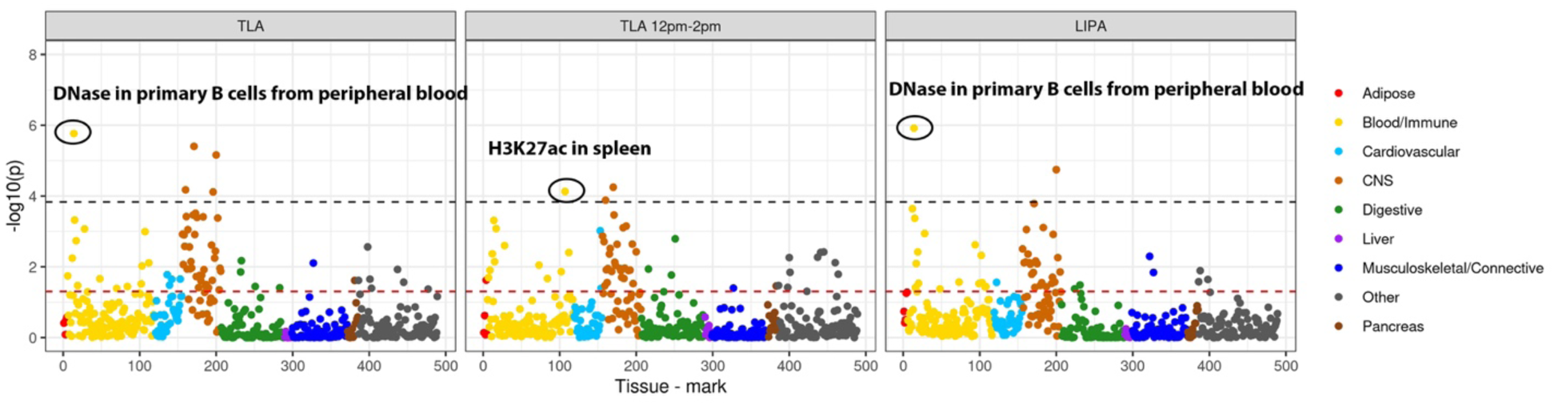
Tissue-specific heritability enrichment p-values for traits with significant enrichment at FDR < 0.05 in blood and immune tissues. The analysis was conducted using tissue/cell type specific stratified LD score regression based on 6 chromatin-based annotations in 111 tissues and cell types described in Finucane et al, Nature Genetics 2018 (PMID: 29632380). Each dot corresponds to an annotation in a tissue or cell type. A complete list of tissue and cell types is provided in Supplementary Table 7 of the above paper. Black line corresponds to FDR < 0.05 (-log(p-value)=3.83) across all combinations of trait, tissue, and histone mark. Red line corresponds to p = 0.05. See Figure S6 for the enrichment p-values for the rest of the traits. CNS: central nervous system.

We further used LD score regression(Bulik-Sullivan et al., 2015a; Bulik-Sullivan et al., 2015b) to explore genetic correlation between PA phenotypes and four broad groups of complex traits and diseases (**Figure S7**). Genetic correlations were identified (FDR < 10%) between PA phenotypes and: (1) neurological, psychiatric and cognitive traits, including Alzheimer’s disease (AD), attention-deficit hyperactivity disorder (ADHD), depressive symptoms, intelligence, and neo-conscientiousness; (2) auto-immune diseases, with the strongest correlation for multiple sclerosis and weaker correlations for Crohn’s disease and primary billary cirrhosis; (3) obesity-related anthropometric traits and (4) cholesterol levels. Most PA traits have negative genetic correlation with obesity-related traits and triglycerides, and positive genetic correlation with HDL cholesterol. The directions of genetic correlation with the other two categories of traits are mixed (**Figure S7**). These results broadly supported our previous results indicating the role of CNS and blood/immune related mechanisms in the genetics of PA traits.

## Discussion

In summary, our study provided novel insights to genetic architecture of physical activity through genome-wide association analysis of an extensive set of accelerometry based PA phenotypes, derived in the UK biobank study, and a series of follow-up genomic analyses. We identified a total of six novel loci, most of which were associated with PA phenotypes not considered in previous studies (Dashti et al., 2019; Doherty et al., 2018; Jones et al., 2019; Klimentidis et al., 2018). Our analysis also identified novel phenotypes associated with the known loci. Further, we provided multiple independent lines of evidence that genetic mechanisms for association for PA involve the blood and immune system.

Compared to the 15 loci identified by the two previous GWASs on accelerometry-based PA (Doherty et al., 2018; Klimentidis et al., 2018), the novel loci we discovered have increased the number of PA susceptibility loci by 33%. Most of the novel loci were connected to the expression of genes, pseudogenes or long non-coding RNAs (lncRNA, **Table 2 and 3**), and *C3orf62* was also supported by evidence of colocalization (**Table S3**). The novel locus indexed by rs9818758 overlaps with the TWAS locus index by *C3orf62*. Though it was unclear how *C3orf62* is involved in PA, two secondary genes in the locus, *ARIH2* and *DAG1* (**Table 3**), appeared to be involved in the following biological processes: *ARIH2* was found to be essential for embryogenesis by regulating the immune system(Lin et al., 2013); *DAG1* was found to play a role in the regeneration of skeletal muscles(Cohn et al., 2002). Both processes appeared relevant for PA. We argue that the two loci above, supported by multiple lines of evidence including GWAS, TWAS, colocalization and gene functions, should be prioritized in follow-up studies. Another two novel loci were connected to pseudogene *RN7SKP16* and *PDXDC2P* of which the function is less clear and may also be worth future investigation. However, Dashti et al found a transcription factor site variant rs915416 to be associated with sleep duration, which is approximately 930Kb away from the transcription start site of *RN7SKP1* and might have potential long range regulatory effects which warrants further study. Among the genes located in known loci, *PBX3* and *RP11-396F22.1* were highlighted by both TWAS and colocalization results. *PBX3* is a member of the pre-B cell leukemia (PBX) family which have extensive roles in early development and some adult processes (Morgan & Pandha, 2020), which could also modulate its association with PA.

The novel phenotypes in this study provided important insights into the genetic architecture of PA, which may have been overlooked by previous GWASs on a small number of phenotypes. The accelerometry-based study by Doherty et al identified the genetic associations with overall activity, sleep duration and sedentary time(Doherty et al., 2018); the study by Klimentidis et al studied the average acceleration and the duration of active states(Klimentidis et al., 2018). Our results found that there can be different genetic architecture for PA during different times of the day, and there can be unique variants that only affect certain PA patterns, like ASTP, LIPA and relative amplitude, but not others (**Table 2**). The heritability and genetic correlation can also vary across different PA phenotypes (**Figures S4 and S7**).

TWAS and tissue-specific heritability enrichment analysis suggested that in addition to the CNS, the blood and immune system could be also associated with PA. This finding was further supported by colocalization, gene-set enrichment and genetic correlation analyses. A previous study(Doherty et al., 2018), which explored enrichment of heritability for PA traits by tissue-specific gene expression patterns, identified potential modulating role of the CNS, adrenal/pancreatic and skeletal muscle tissues. Our study, which used a more extended set of phenotypes and chromatin-state-based annotations, confirmed previous findings and further highlighted the role of the blood and immune system. Though there is lack of studies on the effect of immune functions on PA, previous medical literature has established the effect of PA on immune functions. A study showed that higher PA is associated with elevation of T-regulatory cells and lower risk for autoimmune diseases(Sharif et al., 2018). Multiple studies showed that regular moderately intense PA boost immune functions in older adults and protects against age-related inflammatory disorders(Dhalwani et al., 2016; Duggal, Niemiro, Harridge, Simpson, & Lord, 2019; Vancampfort et al., 2017).Though the direction of causal effect may not be the same as that suggested by genetic analyses, these studies supported broad connections between PA and immune functions. Future studies are needed to better understand the underlying mechanisms and causal directions.

In addition to the blood and immune system, TWAS and enrichment analysis also suggested that the digestive system and endocrine system could be involved in modulating the genetic effects on PA. The literature has also established broad connections between PA and digestive and endocrine tissues. A previous study found that PA has complex effects on gastroinstestinal health(Peters, De Vries, Vanberge-Henegouwen, & Akkermans, 2001): acute strenuous activity may provoke gastrointestinal symptoms while low-intensity activity could have benefits.

Interestingly, three TWAS loci that were significant in digestive tissues were associated with PA phenotypes that are proxies for meal-time activity: *PDXDC2P* with TLA 6am-8am, *RERE* with TLA 6pm-8PM and *KANSL1* with TLA 4pm-6pm (**Table 3**). It was also known that multiple organs in the endocrine system produce hormones that regulate physiological functions of the body, which can have complex bidirectional relationships with PA(Ciloglu et al., 2005; Hawkins et al., 2008; Ennour-Idrissi, Maunsell, & Diorio, 2015; Alessa et al., 2017; Hackney & Saeidi, 2019).

Among the endocrine tissues, thyroid appeared to be modulating the genetic effect of the largest number of loci. Previous studies showed that TWAS lead genes *PBX3*, *Corf62* and *KANSL1* were highly expressed in thyroid (GTEx et al., 2017), and SNPs near *KANSL1* were found to be associated with thyroid-stimulating hormone levels (Teumer et al., 2018). Our TWAS analysis indicated that the genes associated with PA via the blood and immune system tended to also be associated with the digestive and endocrine systems, but do not usually overlap with the genes associated with the CNS. This suggests that the blood and immune, digestive and endocrine systems may be involved in the same broad pathway that affects PA, which is different from that of CNS.

It is noteworthy that the accelerometry-derived PA phenotypes in this study are not limited to exercise, but include a variety of broad-sense activity patterns. In fact, we do find that several phenotypes have strong genetic correlation with sleep, chronotype and other behavioral traits (**Figure S7**). Therefore, when defining novel loci, we have further excluded those variants previously reported to be associated with PA, sleep and circadian rhythm. However, due to the nature of accelerometry, which captures the acceleration of human body, those phenotypes are still essentially and broadly PA phenotypes, though they can indirectly reflect and be related to other traits. Hence we still address all of them by PA phenotypes throughout the paper.

This study has a number of limitations. Though we derived a more extensive set of PA phenotypes than previous studies, information was still lost when collapsing a 7-day continuous times series of wrist accelerometry into 31 PA phenotypes. The ideal approach would be to conduct a GWAS utilizing all the information across the 7 days of accelerometry measurements. Results could outline genetic regulation of a continuous course of PA over time. The current analysis of TLA during 12 non-overlapping two-hour time intervals during the day, indicated that different genetic variants may affect PA during different times of the day (**Tables 2 and 3**). Another limitation is that some of the phenotypes are not directly interpretable. For example, the PCs of log acceleration are less interpretable than other phenotypes, such as TLA and ASTP. However, they do reflect important features of physical activity and warrant further investigations. A potential solution is to obtain proxy measurements that are interpretable and highly correlated with PC scores.

In conclusion, we conducted association studies on a wide range of PA phenotypes and identified 5 novel loci associated with PA. We found that in addition to the CNS, the blood and immune system may also play an important role in the genetic mechanisms of PA, and the digestive and endocrine systems could also be involved in the blood and immune pathway.

## Data Availability

Data supporting the findings of this paper are available upon application to the UK Biobank study. The summary statistics are publicly available via the GWAS Catalog (https://www.ebi.ac.uk/gwas/) under accession numbers GCST90061408-GCST90061434.

## Supplemental Data

Supplemental Data include seven figures and four tables.

## Acknowledgements

The UK Biobank data was accessed via application ID 17712. Research of Drs. Guanghao Qi, Diptavo Dutta and Nilanjan Chatterjee was supported by an R01 grant from the National Human Genome Research Institute [1 R01 HG010480-01].

## Web Resources

UK Biobank: https://www.ukbiobank.ac.uk/

fastGWA software: https://cnsgenomics.com/software/gcta/#fastGWA

FUSION TWAS software: http://gusevlab.org/projects/fusion/

COLOC R package: https://cran.r-project.org/web/packages/coloc/index.html

LD score regression software: https://github.com/bulik/ldsc

LD Hub: http://ldsc.broadinstitute.org/ldhub/

FUMA GWAS: https://fuma.ctglab.nl/

PLINK: https://www.cog-genomics.org/plink/2.0/

Molecular Signatures Database: http://www.broadinstitute.org/msigdb

Open Targets Genetics: https://genetics.opentargets.org/

