## Supplemental Tables for "Genome-wide association studies of 27 accelerometry-derived physical activity measurements identified novel loci and genetic mechanisms"

### Table S1. Traits associated with other variants in the GWAS significant loci (Table 1) but not significantly associated with the lead variant.

| SNP | CHR | BP | Other traits associated with variants tagged by the lead variant[a] |
|---|---|---|---|
| Novel loci | | | |
| rs301799 | 1 | 8489302 | ASTP, Relative amplitude |
| rs9818758 | 3 | 49382925 | TLA 2am-4am |
| 3:131647162_TA_T | 3 | 131647162 | TLA 6am-8am, PC2 |
| Known loci | | | |
| rs2138543 | 12 | 39298423 | Timing of L5, TLA 12am-2am, TLA 2am-4am, L5, Relative amplitude, TLA 6pm-8pm, LIPA, TLA 8pm-10pm, TLA, TLA 10pm-12am, MVPA |
| rs1268539 | 9 | 128195657 | SATP |

[a] Here we use the same significant threshold as in Table 2, which is $p < 5 \times 10^{-8}/19$. Variants that have $r^2 > 0.8$ and are within 500kb of the lead variant are considered tagged. For loci in Table 1 but not in this Table, other tagged SNPs did not have associated traits that are not captured by the lead variant.

**Table S2. All significant tissue-trait pairs in TWAS analysis.** Tissue categories were obtained from Supplementary Table 2 of Finucane et al, Nat Genet 2018, PMID 29632380.

| Gene | CHR | Gene start | Gene end | TWAS p-value | Trait | Tissue | Tissue category |
|---|---|---|---|---|---|---|---|
| RN7SKP16 | 1 | 33802167 | 33802465 | 2.72e-09 (-) | MVPA | Brain Putamen basal ganglia | CNS |
| PDXDC2P | 16 | 70069541 | 70098679 | 3.41e-09 (-) | TLA 6am-8am | Cells EBV-transformed lymphocytes | Blood/Immune |
| PDXDC2P | 16 | 70069541 | 70098679 | 1.81e-09 (-) | TLA 6am-8am | Esophagus Mucosa | Digestive |
| RERE | 1 | 8412457 | 8877702 | 5.28e-09 (-) | TLA 6pm-8pm | Whole Blood | Blood/Immune |
| RERE | 1 | 8412457 | 8877702 | 1.67e-08 (-) | TLA 6pm-8pm | Spleen | Blood/Immune |
| RERE | 1 | 8412457 | 8877702 | 6.5e-09 (-) | TLA 6pm-8pm | Esophagus Mucosa | Digestive |
| RERE | 1 | 8412457 | 8877702 | 8.78e-09 (-) | TLA 6pm-8pm | Skin Not Sun Exposed Suprapubic | Other |
| RP5-1160K1.6 | 1 | 110171118 | 110171939 | 7.68e-10 (-) | PC2 | Brain Putamen basal ganglia | CNS |
| C3orf62 | 3 | 49306219 | 49315263 | 4.94e-09 (+) | Relative amplitude | Esophagus Mucosa | Digestive |
| C3orf62 | 3 | 49306219 | 49315263 | 1.75e-08 (+) | Relative amplitude | Colon Sigmoid | Digestive |
| C3orf62 | 3 | 49306219 | 49315263 | 3.28e-10 (+) | Relative amplitude | Thyroid | Endocrine |
| CTC-467M3.3 | 5 | 87988462 | 87989789 | 1.39e-08 (-) | ASTP | Brain Frontal Cortex BA9 | CNS |
| NMUR2 | 5 | 151771093 | 151812929 | 9.84e-10 (+) | TLA 6pm-8pm | Brain Spinal cord cervical c-1 | CNS |
| NMUR2 | 5 | 151771093 | 151812929 | 8.88e-10 (+) | TLA 6pm-8pm | Prostate | Other |
| PBX3 | 9 | 128509624 | 128729656 | 1.63e-09 (-) | TLA | Cells EBV-transformed lymphocytes | Blood/Immune |
| PBX3 | 9 | 128509624 | 128729656 | 5.59e-10 (-) | TLA | Esophagus Muscularis | Digestive |
| PBX3 | 9 | 128509624 | 128729656 | 6.17e-09 (-) | TLA | Thyroid | Endocrine |
| PBX3 | 9 | 128509624 | 128729656 | 7.96e-10 (-) | TLA | Cells Transformed fibroblasts | Other |
| PBX3 | 9 | 128509624 | 128729656 | 3.09e-09 (-) | TLA | Vagina | Other |
| PBX3 | 9 | 128509624 | 128729656 | 9.91e-10 (-) | LIPA | Cells EBV-transformed lymphocytes | Blood/Immune |
| PBX3 | 9 | 128509624 | 128729656 | 1.59e-08 (-) | LIPA | Esophagus Muscularis | Digestive |
| PBX3 | 9 | 128509624 | 128729656 | 3.04e-10 (-) | LIPA | Thyroid | Endocrine |
| PBX3 | 9 | 128509624 | 128729656 | 3.88e-08 (-) | LIPA | Vagina | Other |
| PBX3 | 9 | 128509624 | 128729656 | 4.36e-09 (-) | SATP | Cells EBV-transformed lymphocytes | Blood/Immune |
| PBX3 | 9 | 128509624 | 128729656 | 7.7e-09 (-) | SATP | Esophagus Muscularis | Digestive |
| PBX3 | 9 | 128509624 | 128729656 | 2.94e-08 (-) | SATP | Thyroid | Endocrine |
| PBX3 | 9 | 128509624 | 128729656 | 2.92e-09 (-) | SATP | Cells Transformed fibroblasts | Other |
| PBX3 | 9 | 128509624 | 128729656 | 7.55e-09 (-) | SATP | Vagina | Other |
| DNAJC1 | 10 | 22045466 | 22292698 | 3.2e-09 (+) | TLA 8am-10am | Brain Anterior cingulate cortex BA24 | CNS |

| | | | | | | | |
|---|---|---|---|---|---|---|---|
| RP11-396F22.1 | 12 | 39300253 | 39303394 | 9.89e-10 (+) | TLA 6am-8am | Brain Frontal Cortex BA9 | CNS |
| RP11-396F22.1 | 12 | 39300253 | 39303394 | 8e-09 (+) | TLA 6am-8am | Brain Anterior cingulate cortex BA24 | CNS |
| RP11-396F22.1 | 12 | 39300253 | 39303394 | 1.03e-08 (+) | TLA 6am-8am | Lung | Other |
| RP11-396F22.1 | 12 | 39300253 | 39303394 | 2.98e-09 (-) | PC2 | Brain Anterior cingulate cortex BA24 | CNS |
| RP11-396F22.1 | 12 | 39300253 | 39303394 | 1.08e-08 (-) | PC2 | Brain Frontal Cortex BA9 | CNS |
| RP11-396F22.1 | 12 | 39300253 | 39303394 | 9.7e-09 (-) | PC2 | Lung | Other |
| RP11-396F22.1 | 12 | 39300253 | 39303394 | 9.51e-09 (-) | Timing of M10 | Brain Anterior cingulate cortex BA24 | CNS |
| RP11-396F22.1 | 12 | 39300253 | 39303394 | 1.4e-08 (-) | Timing of M10 | Brain Frontal Cortex BA9 | CNS |
| FMNL1 | 17 | 43299590 | 43324633 | 2.82e-09 (-) | TLA | Brain Cerebellum | CNS |
| KANSL1 | 17 | 44107282 | 44302733 | 1.83e-09 (+) | TLA | Adipose Subcutaneous | Adipose |
| KANSL1 | 17 | 44107282 | 44302733 | 2.17e-13 (+) | TLA | Cells EBV-transformed lymphocytes | Blood/Immune |
| KANSL1 | 17 | 44107282 | 44302733 | 3.64e-11 (+) | TLA | Whole Blood | Blood/Immune |
| KANSL1 | 17 | 44107282 | 44302733 | 1.44e-08 (+) | TLA | Heart Left Ventricle | Cardiovascular |
| KANSL1 | 17 | 44107282 | 44302733 | 4.94e-11 (+) | TLA | Brain Putamen basal ganglia | CNS |
| KANSL1 | 17 | 44107282 | 44302733 | 1.13e-10 (+) | TLA | Brain Cerebellar Hemisphere | CNS |
| KANSL1 | 17 | 44107282 | 44302733 | 2.09e-10 (+) | TLA | Brain Caudate basal ganglia | CNS |
| KANSL1 | 17 | 44107282 | 44302733 | 5.36e-10 (+) | TLA | Brain Cerebellum | CNS |
| KANSL1 | 17 | 44107282 | 44302733 | 1.4e-08 (+) | TLA | Brain Substantia nigra | CNS |
| KANSL1 | 17 | 44107282 | 44302733 | 3.87e-08 (+) | TLA | Brain Cortex | CNS |
| KANSL1 | 17 | 44107282 | 44302733 | 5.44e-11 (+) | TLA | Small Intestine Terminal Ileum | Digestive |
| KANSL1 | 17 | 44107282 | 44302733 | 9.8e-11 (+) | TLA | Esophagus Mucosa | Digestive |
| KANSL1 | 17 | 44107282 | 44302733 | 7.22e-09 (+) | TLA | Colon Sigmoid | Digestive |
| KANSL1 | 17 | 44107282 | 44302733 | 2.85e-08 (+) | TLA | Colon Transverse | Digestive |
| KANSL1 | 17 | 44107282 | 44302733 | 9.15e-08 (+) | TLA | Esophagus Gastroesophageal Junction | Digestive |
| KANSL1 | 17 | 44107282 | 44302733 | 6.63e-10 (+) | TLA | Pituitary | Endocrine |
| KANSL1 | 17 | 44107282 | 44302733 | 8.31e-11 (+) | TLA | Muscle Skeletal | Musculoskeletal/connective |
| KANSL1 | 17 | 44107282 | 44302733 | 1.96e-12 (+) | TLA | Cells Transformed fibroblasts | Other |
| KANSL1 | 17 | 44107282 | 44302733 | 3.33e-12 (+) | TLA | Uterus | Other |
| KANSL1 | 17 | 44107282 | 44302733 | 3.62e-11 (+) | TLA | Skin Sun Exposed Lower leg | Other |
| KANSL1 | 17 | 44107282 | 44302733 | 7e-10 (+) | TLA | Lung | Other |
| KANSL1 | 17 | 44107282 | 44302733 | 1.47e-09 (+) | TLA | Minor Salivary Gland | Other |
| KANSL1 | 17 | 44107282 | 44302733 | 3.75e-09 (+) | TLA | Skin Not Sun Exposed Suprapubic | Other |
| KANSL1 | 17 | 44107282 | 44302733 | 1.49e-08 (+) | TLA | Ovary | Other |

| | | | | | | | |
|---|---|---|---|---|---|---|---|
| KANSL1 | 17 | 44107282 | 44302733 | 9.13e-08 (+) | TLA | Vagina | Other |
| KANSL1 | 17 | 44107282 | 44302733 | 3.09e-08 (+) | TLA 4pm-6pm | Cells EBV-transformed lymphocytes | Blood/Immune |
| KANSL1 | 17 | 44107282 | 44302733 | 4.7e-08 (+) | TLA 4pm-6pm | Small Intestine Terminal Ileum | Digestive |
| KANSL1 | 17 | 44107282 | 44302733 | 7e-09 (+) | TLA 4pm-6pm | Uterus | Other |
| KANSL1 | 17 | 44107282 | 44302733 | 1.03e-08 (+) | TLA 4pm-6pm | Skin Sun Exposed Lower leg | Other |
| KANSL1 | 17 | 44107282 | 44302733 | 8.65e-08 (+) | TLA 4pm-6pm | Cells Transformed fibroblasts | Other |
| KANSL1 | 17 | 44107282 | 44302733 | 1.08e-09 (+) | MVPA | Cells EBV-transformed lymphocytes | Blood/Immune |
| KANSL1 | 17 | 44107282 | 44302733 | 5.14e-09 (+) | MVPA | Brain Putamen basal ganglia | CNS |
| KANSL1 | 17 | 44107282 | 44302733 | 7.5e-08 (+) | MVPA | Brain Caudate basal ganglia | CNS |
| KANSL1 | 17 | 44107282 | 44302733 | 9.25e-08 (+) | MVPA | Brain Cerebellar Hemisphere | CNS |
| KANSL1 | 17 | 44107282 | 44302733 | 9.15e-09 (+) | MVPA | Small Intestine Terminal Ileum | Digestive |
| KANSL1 | 17 | 44107282 | 44302733 | 5.67e-08 (+) | MVPA | Esophagus Mucosa | Digestive |
| KANSL1 | 17 | 44107282 | 44302733 | 9.82e-09 (+) | MVPA | Muscle Skeletal | Musculoskeletal/connective |
| KANSL1 | 17 | 44107282 | 44302733 | 2.88e-09 (+) | MVPA | Cells Transformed fibroblasts | Other |
| KANSL1 | 17 | 44107282 | 44302733 | 6.48e-09 (+) | MVPA | Uterus | Other |
| KANSL1 | 17 | 44107282 | 44302733 | 5.74e-08 (+) | MVPA | Skin Sun Exposed Lower leg | Other |
| ZNF846 | 19 | 9862669 | 9903856 | 2.22e-10 (-) | TLA | Nerve Tibial | Other |
| JUND | 19 | 18390563 | 18392432 | 3.18e-09 (+) | SATP | Uterus | Other |
| LINC00634 | 22 | 42348169 | 42354937 | 4.85e-11 (-) | TLA | Brain Cerebellar Hemisphere | CNS |

**Table S3. Nine loci associated with physical activity in Table 2 colocalize with eQTL signals.** We conducted colocalization analysis between the GWAS signal for the most significant trait associated with the locus and eQTL signal for the expression of nearby genes. The analysis was perfomed using COLOC and eQTL summary statistics are from GTEx v7. The locus numbers are consistent with those in Table 2. Tissue categories were obtained from Supplementary Table 2 of Finucane et al, Nat Genet 2018, PMID 29632380.

| Gene | Tissue | Tissue category | PP4 |
|---|---|---|---|
| **Locus #1. rs301799 (chr1:8489302). Most significantly associated trait: TLA 6pm-8pm.** | | | |
| RERE† | Adipose Visceral Omentum | Adipose | 0.924 |
| | Spleen | Blood/Immune | 0.983 |
| | Whole Blood | Blood/Immune | 0.987 |
| | Esophagus Mucosa | Digestive | 0.991 |
| | Thyroid | Endocrine | 0.877 |
| | Lung | Other | 0.966 |
| | Pancreas | Other | 0.968 |
| | Skin Not Sun Exposed Suprapubic | Other | 0.991 |
| | Skin Sun Exposed Lower leg | Other | 0.922 |
| RP5-1115A15.1† | Adipose Subcutaneous | Adipose | 0.991 |
| | Adipose Visceral Omentum | Adipose | 0.975 |
| | Cells EBV-transformed lymphocytes | Blood/Immune | 0.931 |
| | Artery Tibial | Cardiovascular | 0.986 |
| | Esophagus Mucosa | Digestive | 0.984 |
| | Esophagus Muscularis | Digestive | 0.923 |
| | Thyroid | Endocrine | 0.991 |
| | Muscle Skeletal | Musculoskeletal/connective | 0.989 |
| | Breast Mammary Tissue | Other | 0.977 |
| | Lung | Other | 0.975 |
| | Nerve Tibial | Other | 0.975 |
| | Ovary | Other | 0.94 |
| | Pancreas | Other | 0.991 |
| | Skin Not Sun Exposed Suprapubic | Other | 0.992 |
| | Skin Sun Exposed Lower leg | Other | 0.957 |

| |
|---|
| **Locus #2. rs3836464 (chr3:10454772). Most significantly associated trait: ASTP.** |

| | | | |
|---|---|---|---|
| ARPC4 | Artery Tibial | Cardiovascular | 0.969 |
| ATP2B2 | Testis | Endocrine | 0.955 |

| | | | |
|---|---|---|---|
| **Locus #3. rs9818758 (chr3:49382925). Most significantly associated trait: relative amplitude.** | | | |
| C3orf62† | Thyroid | Endocrine | 0.823 |
| | Cells Transformed fibroblasts | Other | 0.824 |
| CCDC36 | Testis | Endocrine | 0.904 |
| DALRD3 | Skin Not Sun Exposed Suprapubic | Other | 0.918 |
| | Skin Sun Exposed Lower leg | Other | 0.877 |
| GPX1 | Nerve Tibial | Other | 0.887 |
| HYAL3 | Adipose Visceral Omentum | Adipose | 0.982 |
| | Artery Tibial | Cardiovascular | 0.913 |
| | Heart Left Ventricle | Cardiovascular | 0.973 |
| | Esophagus Mucosa | Digestive | 0.941 |
| | Esophagus Muscularis | Digestive | 0.811 |
| | Thyroid | Endocrine | 0.855 |
| | Breast Mammary Tissue | Other | 0.944 |
| | Skin Not Sun Exposed Suprapubic | Other | 0.82 |
| IP6K2 | Artery Aorta | Cardiovascular | 0.877 |
| KLHDC8B | Artery Tibial | Cardiovascular | 0.872 |
| | Esophagus Mucosa | Digestive | 0.823 |
| MST1 | Adipose Subcutaneous | Adipose | 0.924 |
| | Colon Sigmoid | Digestive | 0.938 |
| | Colon Transverse | Digestive | 0.896 |
| | Esophagus Mucosa | Digestive | 0.936 |
| | Adrenal Gland | Endocrine | 0.854 |
| | Cells Transformed fibroblasts | Other | 0.929 |
| | Lung | Other | 0.915 |
| PRKAR2A | Skin Sun Exposed Lower leg | Other | 0.884 |
| RBM5 | Esophagus Mucosa | Digestive | 0.849 |
| RBM6 | Pancreas | Other | 0.834 |
| SEMA3F | Nerve Tibial | Other | 0.87 |

| | | | |
|---|---|---|---|
| SHISA5 | Brain Cerebellum | CNS | 0.87 |
| SLC26A6 | Thyroid | Endocrine | 0.928 |
| | Liver | Liver | 0.831 |
| | Muscle Skeletal | Musculoskeletal/connective | 0.869 |
| | Lung | Other | 0.906 |
| TMEM89 | Artery Tibial | Cardiovascular | 0.914 |
| TREX1 | Esophagus Muscularis | Digestive | 0.804 |
| USP19 | Artery Aorta | Cardiovascular | 0.874 |
| | Adrenal Gland | Endocrine | 0.808 |
| WDR6 | Thyroid | Endocrine | 0.915 |
| | Skin Not Sun Exposed Suprapubic | Other | 0.804 |
| | Skin Sun Exposed Lower leg | Other | 0.816 |
| **Locus #5. rs2138543 (chr12:39298423). Most significantly associated trait: TLA 6am-8am.** | | | |
| RP11-396F22.1† | Colon Transverse | Digestive | 0.869 |
| | Lung | Other | 0.94 |
| | Nerve Tibial | Other | 0.966 |
| **Locus #6. rs1144566 (chr1:182569626). Most significantly associated trait: Timing of L5.** | | | |
| NMNAT2 | Adipose Visceral Omentum | Adipose | 0.801 |
| **Locus #12. rs1268539 (chr9:128195657). Most significantly associated trait: TLA.** | | | |
| GAPVD1 | Ovary | Other | 0.924 |
| PBX3† | Heart Atrial Appendage | Cardiovascular | 0.829 |
| | Heart Left Ventricle | Cardiovascular | 0.934 |
| | Esophagus Gastroesophageal Junction | Digestive | 0.936 |
| | Esophagus Muscularis | Digestive | 0.959 |
| **Locus #13. rs564819152 (chr10:21820650). Most significantly associated trait: TLA 8am-10am.** | | | |
| CASC10† | Skin Sun Exposed Lower leg | Other | 0.924 |
| MLLT10 | Skin Sun Exposed Lower leg | Other | 0.971 |

| | | | |
|---|---|---|---|
| **Locus #15. rs2532402 (chr17:44304130). Most significantly associated trait: TLA.** | | | |
| DCAKD | Thyroid | Endocrine | 0.826 |
| NMT1 | Thyroid | Endocrine | 0.918 |
| | Cells Transformed fibroblasts | Other | 0.903 |

| | | | |
|---|---|---|---|
| **Locus #16. rs3837946 (chr19:9955920). Most significantly associated trait: TLA.** | | | |
| CTD-2623N2.11 | Brain Frontal Cortex BA9 | CNS | 0.83 |
| | Brain Putamen basal ganglia | CNS | 0.859 |
| PIN1 | Colon Sigmoid | Digestive | 0.825 |
| ZNF266 | Spleen | Blood/Immune | 0.968 |
| | Adrenal Gland | Endocrine | 0.929 |
| | Pancreas | Other | 0.864 |

† Genes with significant TWAS associations (either lead or secondary gene in Table 2).

**Table S4. Validation of potential novel loci in Open Targets Genetics (OTG).** The goal of this analysis is to ensure novel loci do not have associations with related traits not catalogued in Table 2. We search in OTG for the lead variants of 4 potential novel loci and report (1) associations with traits whose names include the following keywords: accelerometry, physical, exercise, sleep, nap, circadian and chronotype; (2) previously reported GWAS lead variants for these traits that are in linkage disequilibrium ($r^2$>0.5) with the lead variants of our potential novel loci. As a results, locus indexed by rs301799 is no longer considered novel due to associations with daytime napping and sleep duration.

| Lead variant | Trait | P value | OTG study ID |
|---|---|---|---|
| **Associations with lead variant** | | | |
| rs301799 | Morning/evening person (chronotype) | 1.90E-03 | NEALE2_1180 |
| | Strenuous sports or other exercises | 3.40E-04 | GCST006100 |
| | Vigorous physical activity | 7.30E-03 | GCST006098 |
| | Moderate to vigorous physical activity levels | 5.00E-02 | GCST006097 |
| | Accelerometer-based physical activity measurement (average acceleration) | 1.70E-05 | GCST006099 |
| | **Nap during day** | **2.40E-07** | **NEALE2_1190** |
| | Sleep duration | 1.70E-02 | GCST003839 |
| | Other exercises (eg: swimming, cycling, keep fit, bowling) \| types of physical activity in last 4 weeks | 2.80E-02 | NEALE2_6164_2 |
| | Light diy (eg: pruning, watering the lawn) \| types of physical activity in last 4 weeks | 4.40E-02 | NEALE2_6164_4 |
| | **Sleep duration** | **8.10E-07** | **NEALE2_1160** |
| rs3836464 | Sleep duration | 8.80E-03 | GCST003839 |
| | Number of days/week of moderate physical activity 10+ minutes | 1.90E-02 | NEALE2_884 |
| rs9818758 | Strenuous sports or other exercises | 1.50E-05 | GCST006100 |
| | Accelerometer-based physical activity measurement (average acceleration) | 4.30E-03 | GCST006099 |
| | Did your sleep change? | 1.60E-02 | NEALE2_20532 |
| | Strenuous sports \| types of physical activity in last 4 weeks | 1.40E-02 | NEALE2_6164_3 |
| | Other exercises (eg: swimming, cycling, keep fit, bowling) \| types of physical activity in last 4 weeks | 7.80E-04 | NEALE2_6164_2 |
| | None of the above \| types of physical activity in last 4 weeks | 4.80E-02 | NEALE2_6164_100 |
| | Chest pain felt during physical activity | 3.70E-02 | NEALE2_6015 |
| | Nap during day | 2.00E-02 | NEALE2_1190 |
| | Job involves heavy manual or physical work | 2.30E-04 | NEALE2_816 |
| | Sleeping too much | 3.20E-02 | NEALE2_20534 |
| 3:131647162_TA_T | Sleep duration (undersleepers) | 4.40E-02 | GCST006686 |
| (Not present in OTG, used | Number of days/week of moderate physical activity 10+ minutes | 3.70E-02 | NEALE2_884 |
| proxy rs1225043) | Sleep duration | 1.70E-02 | GCST003839 |
| | Sleep disorders | 3.50E-03 | SAIGE_327 |
| | Daytime dozing / sleeping (narcolepsy) | 7.30E-03 | NEALE2_1220 |
| | Sleep duration | 1.40E-03 | NEALE2_1160 |
| | None of the above \| types of physical activity in last 4 weeks | 3.90E-02 | NEALE2_6164_100 |
| | Sleep apnea | 2.60E-03 | SAIGE_327_3 |
| **Other GWAS lead variants in LD with lead variant of potential novel loci** | | | |
| rs301817 | **Daytime nap** | **6.00E-13** | **GCST011494** |
| (In LD with rs301799, $r^2$=0.99) | | | |
