## Supplemental Figures for "Genome-wide association studies of 27 accelerometry-derived physical activity measurements identified novel loci and genetic mechanisms"

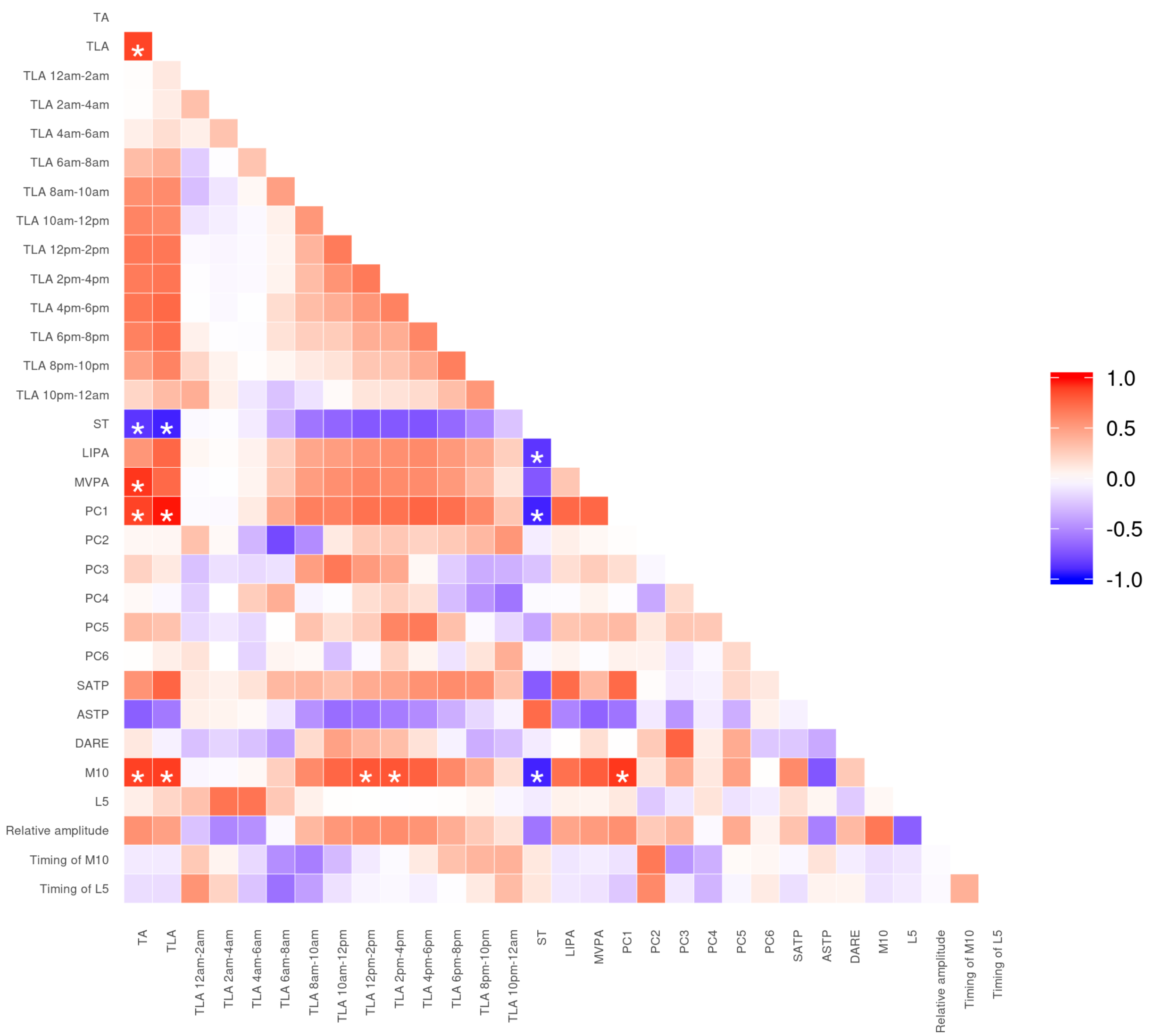


**Figure S1. Correlation matrix for 31 physical activity phenotypes.** Correlation coefficients > 0.8 or < -0.8 are marked with *.

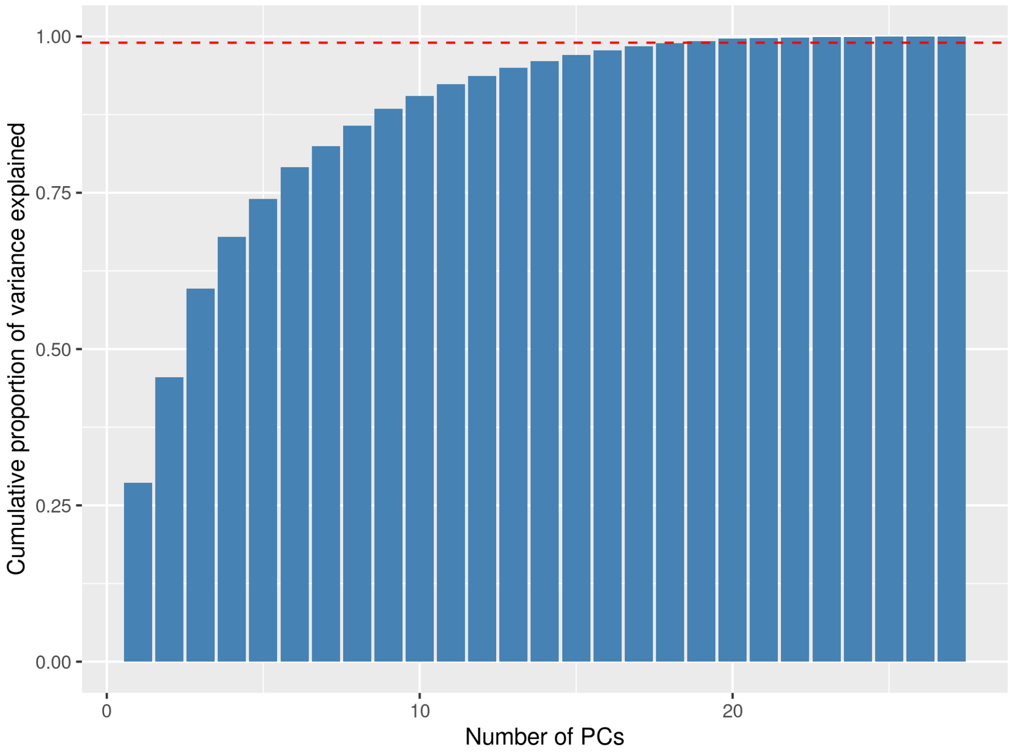


**Figure S2. Number of phenotypic PCs and cumulative proportion of variance explained.** At least 19 PCs are needed to explain 99% of the variance (red dashed line).

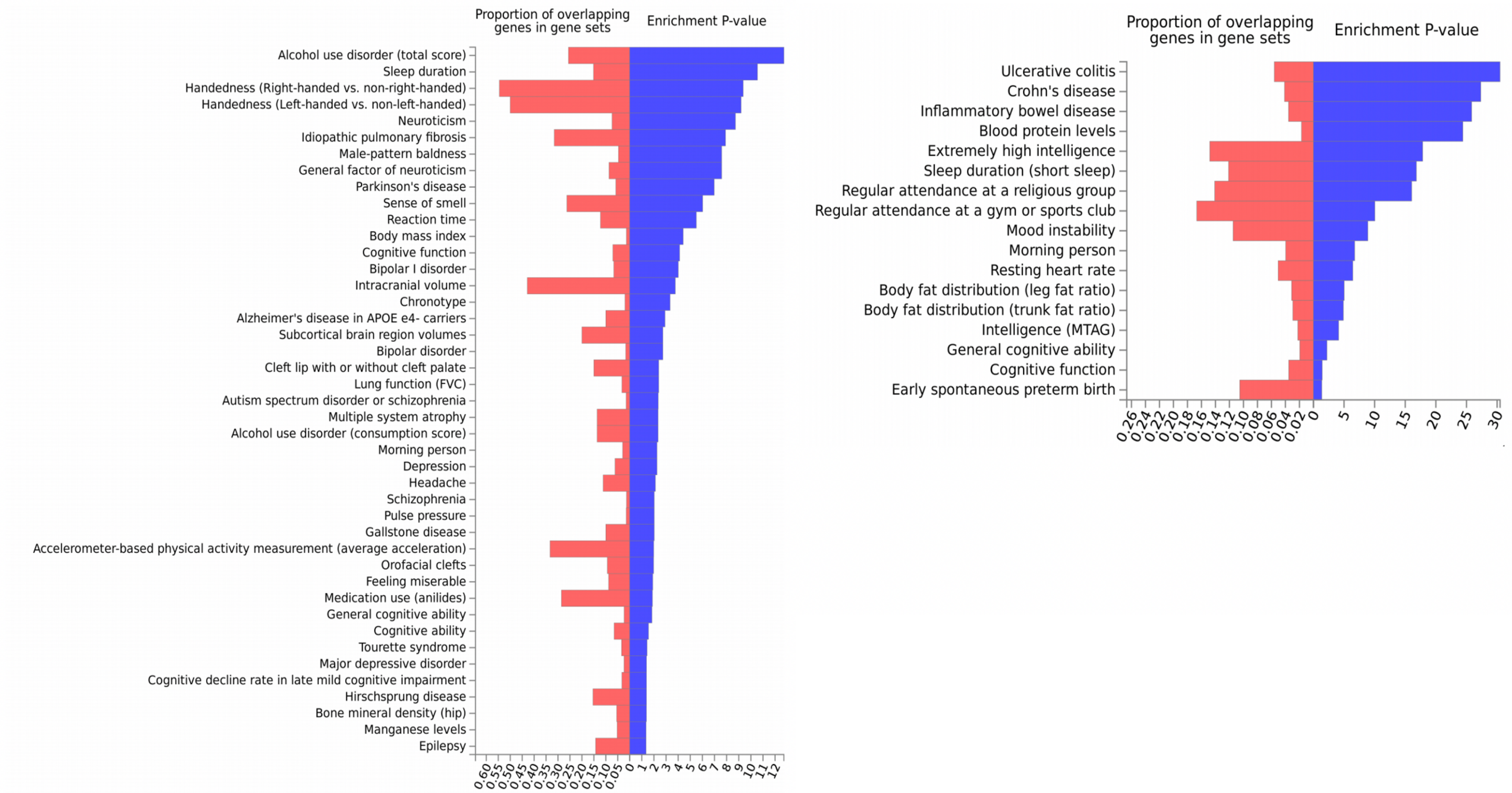


**Figure S3. Gene-set enrichment analysis for the significant genes in TWAS analysis for (A) TLA and (B) relative amplitude.** We use TLA and relative amplitude as examples because they capture two most important patterns of PA: total volume of activity and sleep quality. Genes previously reported to be associated to different traits and diseases as curated in the GWAS catalog were defined as gene-sets (see Methods). The genes associated to (A) TLA and (B) relative amplitude (p-value $< 2.5 \times 10^{-6}$) in transcriptome-wide analysis were checked for enriched overlap with the gene-sets curated from GWAS catalog for each trait (additionally reported in Molecular Signatures database).

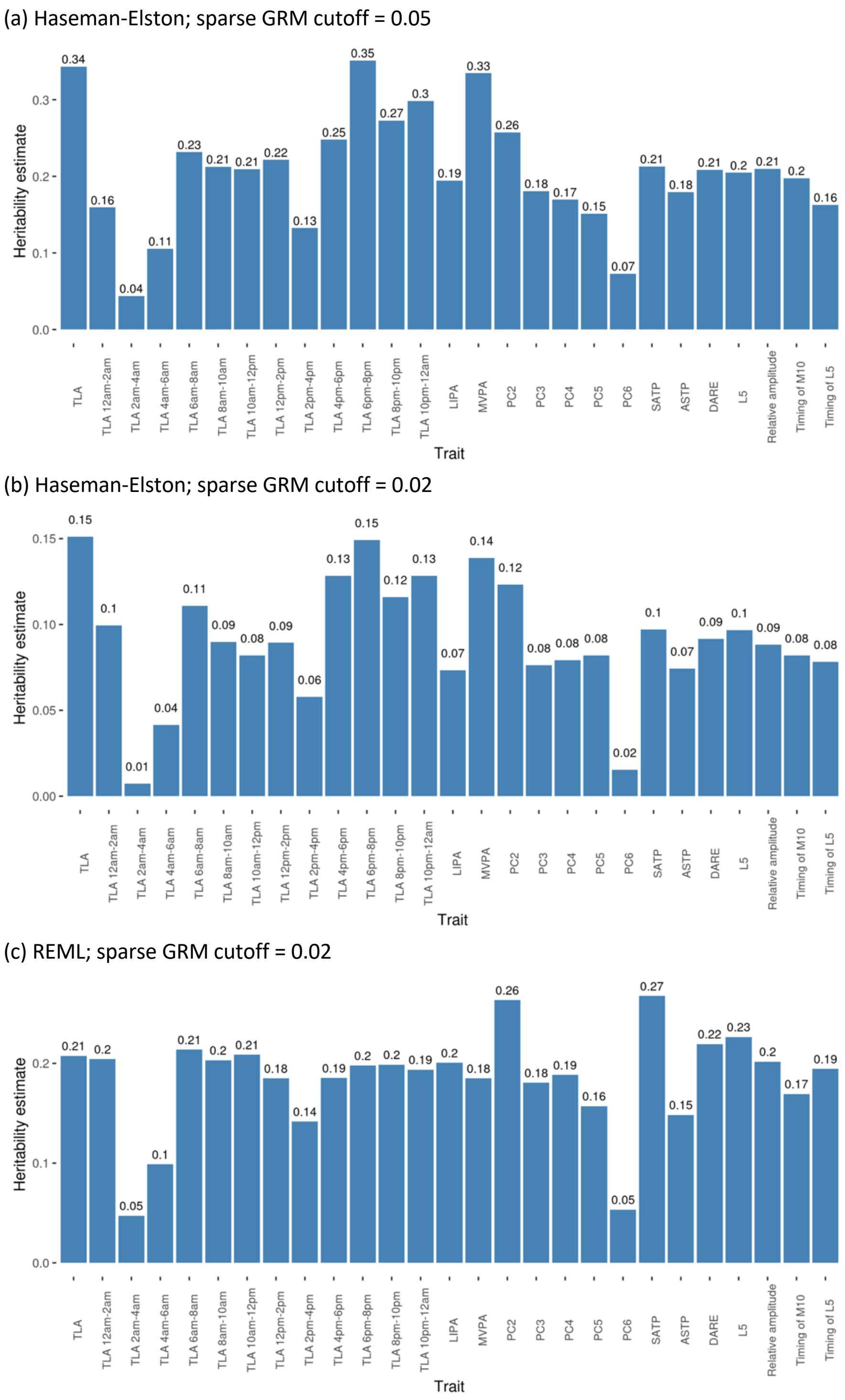


**Figure S4. Estimates of heritability of 27 physical activity traits.** a) Haseman-Elston regression with sparse genetic relationship matrix (GRM) cutoff at 0.05: correlations that are < 0.05 in the GRM are reduced to zero, as is in our fastGWA analysis and recommended by the fastGWA paper. b) Haseman-Elston regression with GRM cutoff at 0.02: correlations that are < 0.02 in the GRM are reduced to zero. c) Restricted maximum likelihood (REML) with GRM cutoff at 0.02.

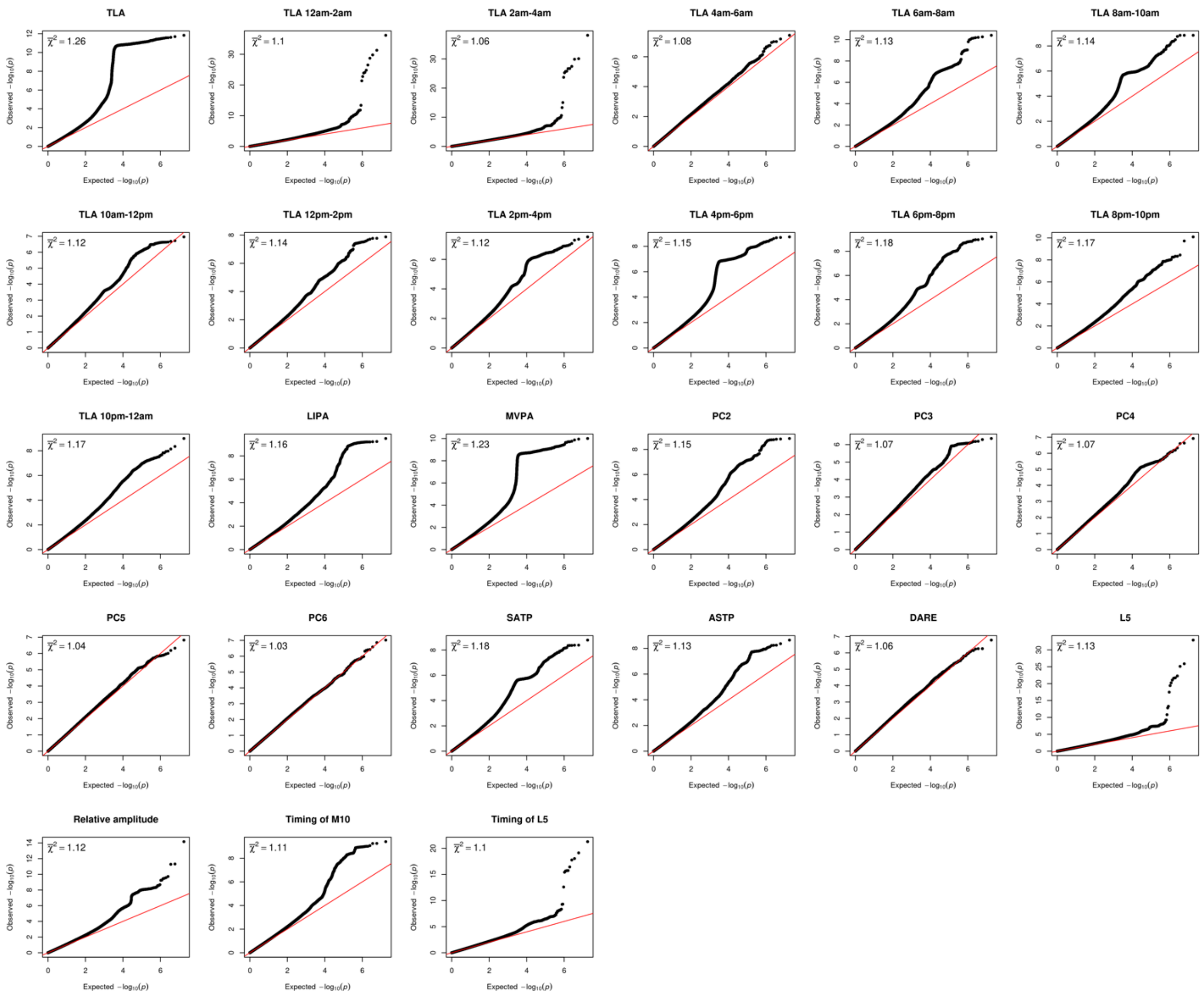


**Figure S5. QQ plot for the p-values from fastGWA analysis for 27 activity phenotypes.** The average $\chi^2$ statistic is annotated at the top left corner of each panel.

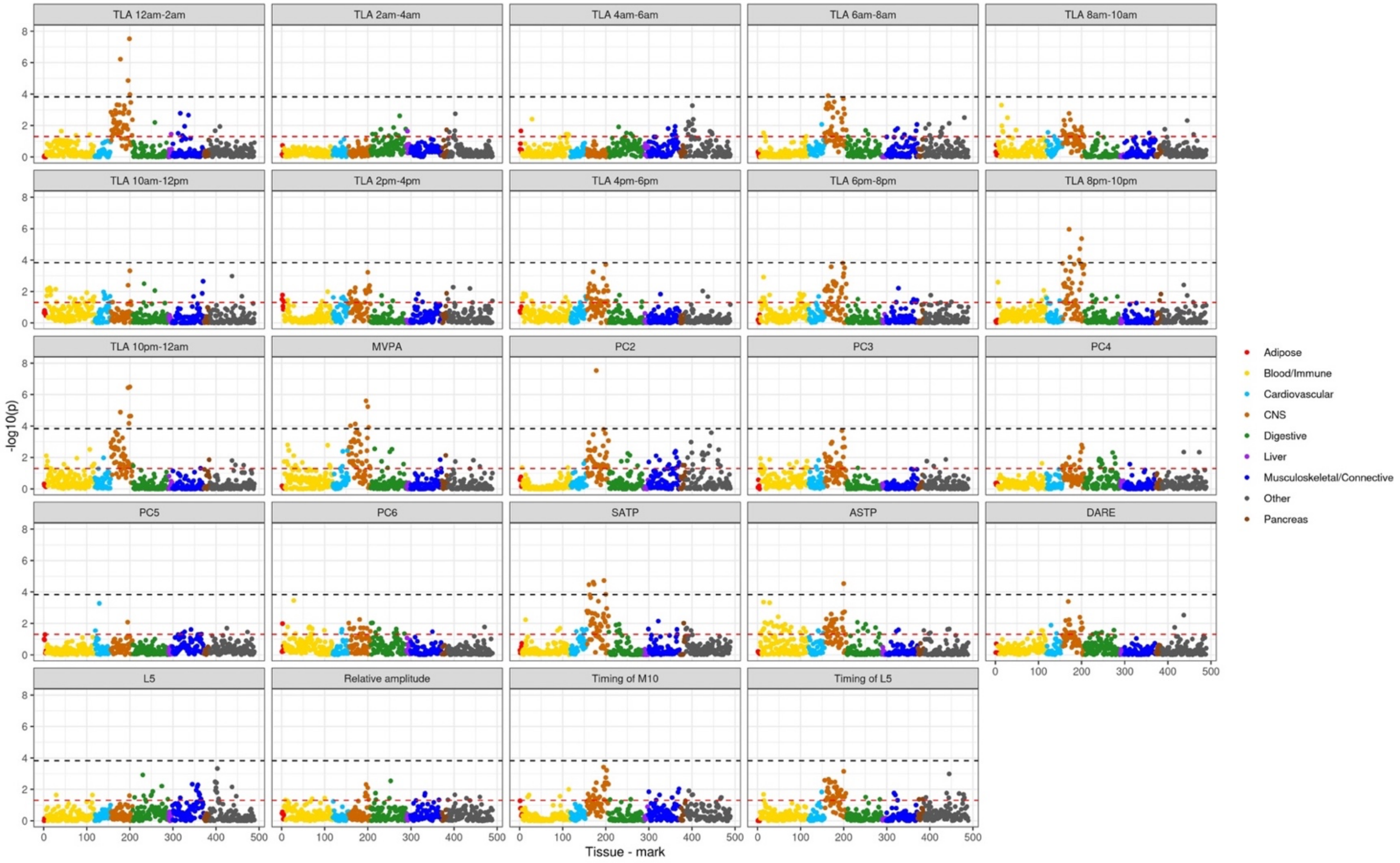


**Figure S6. Tissue-specific heritability enrichment for traits not included in Figure 3**. The analysis was conducted using tissue/cell type specific stratified LD score regression (Finucane et al, Nat Genet 2018, PMID 29632380) based on 6 chromatin-based annotations in 111 tissues and cell types. A complete list of tissue and cell types is provided in Supplementary Table 7 of the above paper. Black line corresponds to FDR < 0.05 (-log(p-value)=3.83) across all combinations of trait, tissue, and histone mark. Red line corresponds to p = 0.05. The analysis was conducted using tissue/cell type specific stratified LD score regression (Finucane et al, 2018).

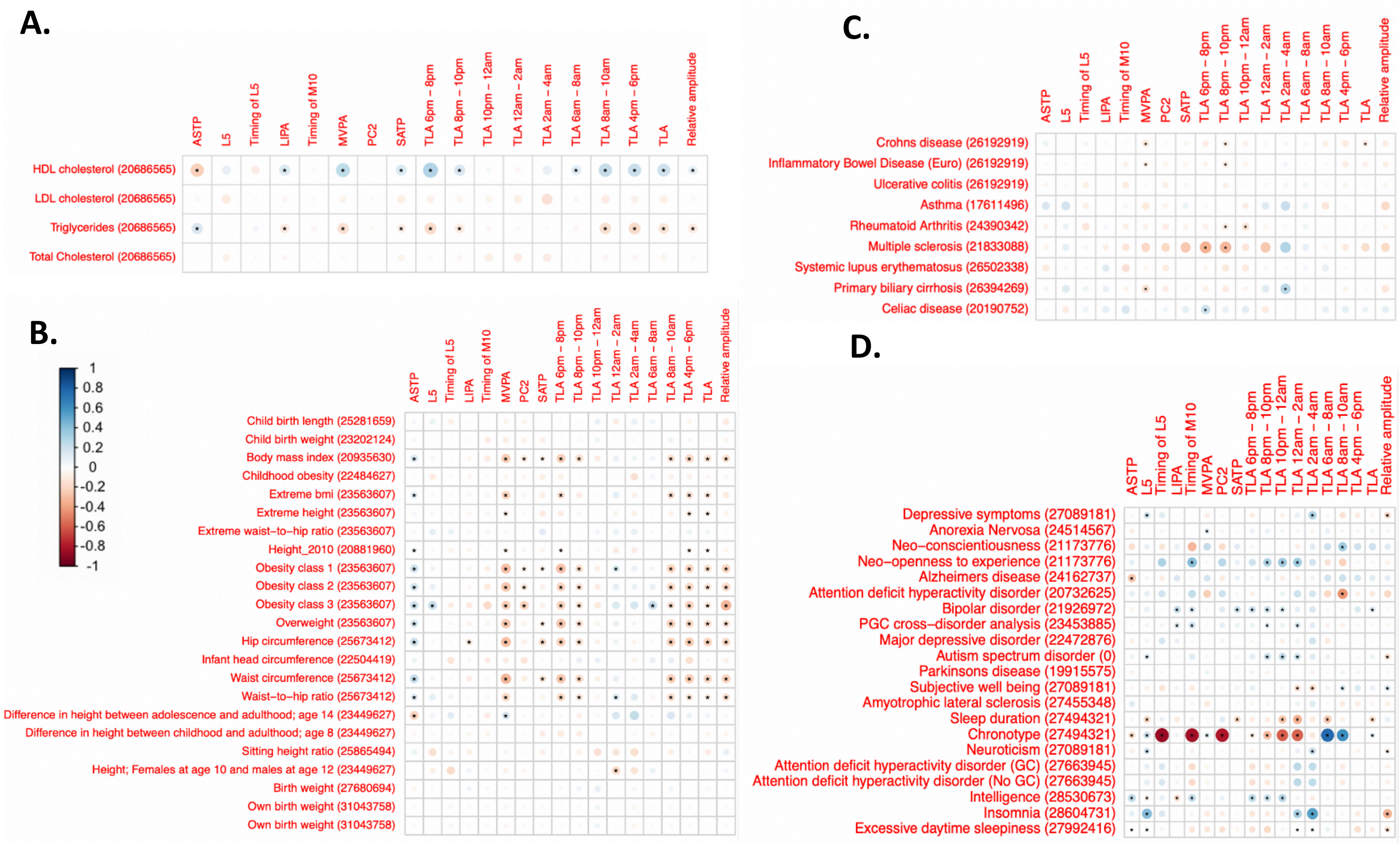


**Figure S7. Genetic correlation of 18 PA traits with other traits: (A) cholesterol levels (B) anthropometric traits (C) autoimmune diseases and (D) miscellaneous traits including psychiatric, neurological, cognitive and personality traits**. The PubMed ID's for the corresponding for the GWAS summary statistics are in parentheses. The size of the circle and the darkness of the color represents the genetic correlation value. The asterisk sign represents if the corresponding genetic correlation was significant at an FDR correction threshold of 10% (corrected within the respective category). For genetic correlation value and p-values across all 237 traits analyzed, see Table S4.
